# Mathematical modeling and adequate environmental sampling plans are essential for the public health assessment of COVID-19 pandemics : development of a monitoring indicator for SARS-CoV-2 in wastewater

**DOI:** 10.1101/2021.09.01.21262877

**Authors:** Nicolas Cluzel, Marie Courbariaux, Siyun Wang, Laurent Moulin, Sébastien Wurtzer, Isabelle Bertrand, Karine Laurent, Patrick Monfort, Obépine consortium, Soizick Le Guyader, Mickaël Boni, Jean-Marie Mouchel, Vincent Maréchal, Grégory Nuel, Yvon Maday

**Affiliations:** Sorbonne Université, Maison des Modélisations Ingénieries et Technologies (SUMMIT), 75005 Paris, France; Eau de Paris, Département de Recherche, Développement et Qualité de l’Eau, 33 avenue Jean Jaurès, F-94200 Ivry sur Seine, France; Université de Lorraine, CNRS, LCPME, F-54000, Nancy, France; HydroSciences Montpellier, UMR 5151, Université de Montpellier, CNRS, IRD, F-34093 Montpellier, France; Ifremer, laboratoire de Microbiologie, SG2M/LSEM, BP 21105, 44311 Nantes, France; Institut de Recherche Biomédicale des Armées, Microbiology & Infectious diseases, Virology unit, 1 place Valérie André, F-91220 Brétigny-sur-Orge, France; Sorbonne Université, CNRS, EPHE, UMR 7619 Metis, e-LTER Zone Atelier Seine, F-75005 Paris, France; Sorbonne Université, INSERM, Centre de Recherche Saint-Antoine, F-75012, Paris, France; Stochastics and Biology Group, Probability and Statistics (LPSM, CNRS 8001), Sorbonne University, Campus Pierre et Marie Curie, 4 Place Jussieu, 75005, Paris, France; Sorbonne Université, CNRS, Université de Paris, Laboratoire Jacques-Louis Lions (LJLL), F-75005 Paris, France

**Keywords:** Wastewater-Based Epidemiology (WBE), Severe Acute Respiratory Syndrome Coronavirus 2 (SARS-CoV-2), Coronavirus Infectious Disease 19 (COVID-19), Mathematical modeling, Correlation, Sampling frequency

## Abstract

Since many infected people experience no or few symptoms, the SARS-CoV-2 epidemic is frequently monitored through massive virus testing of the population, an approach that may be biased and may be difficult to sustain in low-income countries. Since SARS-CoV-2 RNA can be detected in stool samples, quantifying SARS-CoV-2 genome by RT-qPCR in WWTPs^1^ has been proposed as an alternative tool to monitor virus circulation among human populations. However, measuring SARS-CoV-2 viral load in WWTPs can be affected by many experimental and environmental factors. To circumvent these limits, we propose here a novel indicator WWI^2^ that partly reduces and corrects the noise associated with the SARS-CoV-2 genome quantification in wastewater. This method has been successfully applied in the context of Obepine, a French national network that has been quantifying SARS-CoV-2 genome in a representative sample of French WWTPs since March 5th 2020. On August 26th, 2021, 168 WWTPs were monitored twice a week in the metropolitan and overseas territories of France. We detail the process of elaboration of this indicator, show that it is strongly correlated to the incidence rate and that the optimal time lag between these two signals is only a few days, making our indicator an efficient complement or even a credible alternative to the incidence rate. This alternative approach may be especially important to evaluate SARS-CoV-2 dynamics in human populations when the testing rate is low.

**Figure 1:**
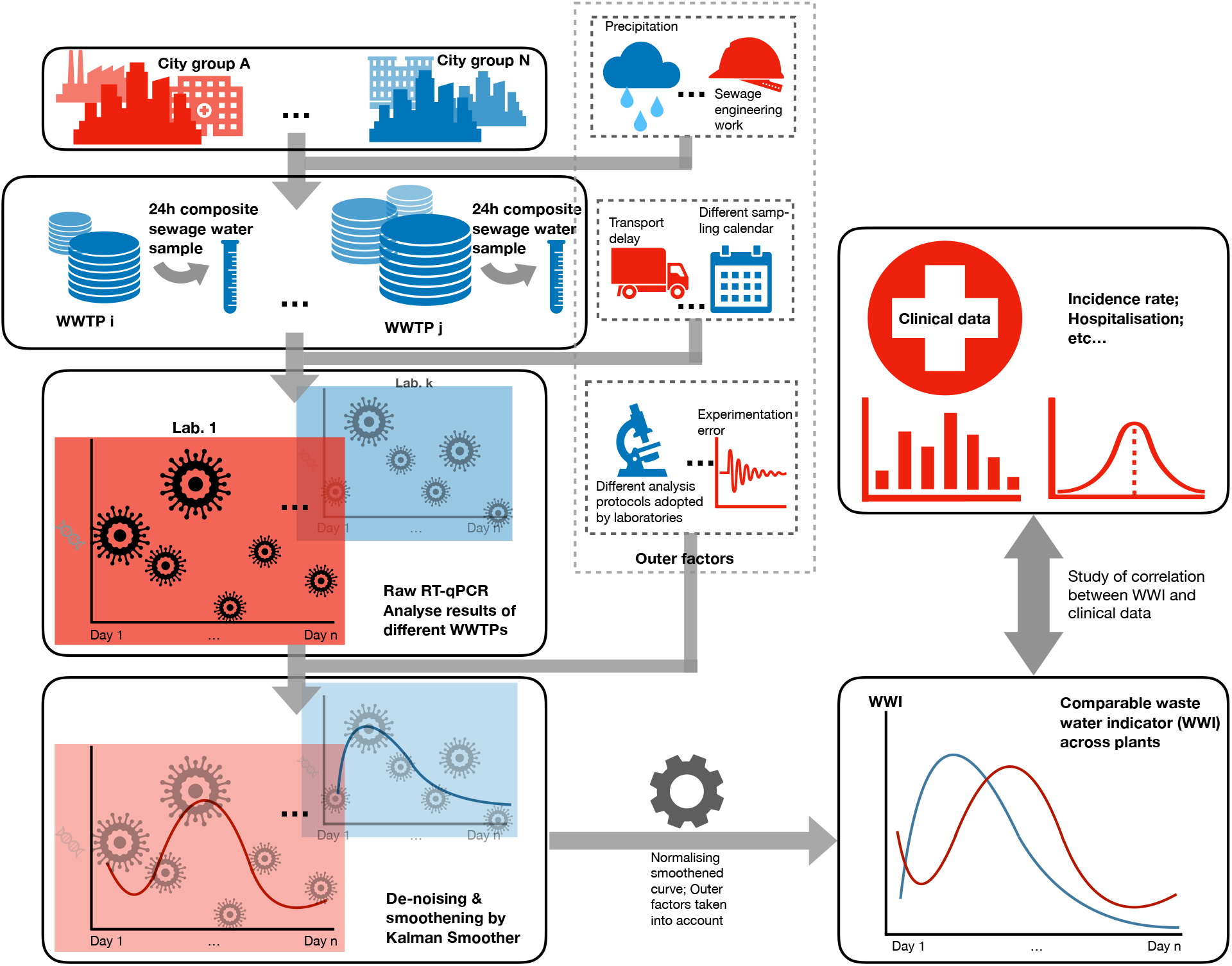
Graphical abstract.

## 1 Introduction

The SARS-CoV-2 pandemic has affected 214 million people worldwide and resulted in more than 6.6 million confirmed cases in France as of August 26th 2021. However, these figures underestimate the total number of infected people. Indeed, many asymptomatic virus carriers are not detected, except during random testing or when they are tested prior to travelling or as contact cases [13, 14]. Moreover, infected people with mild symptoms who do not seek medical assistance will not be screened either. Finally, massive individual testing may vary depending on the epidemiological situation and is economically difficult to sustain, particularly in low income countries.

Several studies have demonstrated the value of wastewater-based epidemiology for monitoring SARS-CoV-2 genome shedding in WWTPs as a putative surrogate or complementary approach to classical epidemiological indicators [1, 9, 11, 12]. However, SARS-CoV-2 genome quantification in wastewater is subject to a number of shortcomings that must be corrected before such monitoring can be deployed on a large scale. These notably include (i) the intralaboratory variability, i.e. the repeatability error on measurements from the same sample and (ii) the inter-laboratory variability, i.e. the difference in genomic units per liter of effluent evaluated by two different laboratories for identical samples even when using similar procedures. (iii) Finally, the specificity of each wastewater network (unitary or separative), its topography, the proportion of industries and the characterization of their discharges are also criteria of variability that must be taken into account to be able to compare the evolution of the epidemics at a regional scale or to deduce the trend nation-wide. The aforementionned variabilities must be corrected if the final purpose is a national monitoring network involving several laboratories, different protocols and many WWTPs. We propose herein an original design of a uniform indicator, WWI, that monitors viral load level in wastewater along time and that takes into account the above-mentioned variabilities. Its performance was assessed on 24 WWTPs followed by the Obepine network, a French national program that has been quantifying SARS-CoV-2 on some of the most important French WWTPs since March 3rd 2020. On August 26th 2021, 168 WWTPs were monitored twice a week. The WWI was compared to local case incidence on different EPCIs^3^. The robustness of this indicator to flow variations linked to various phenomena (rainfalls, civil engineering on the network imposing the detour of the watershed towards other plants, etc.) was estimated. Finally, we compared this indicator to the local incidence rate in order to estimate the correlation, the time lag between these two signals as well as the capacity of the WWI to anticipate major epidemiological changes (increased viral circulation, reduced circulation in response to governmental measures for example). This study focused on the peak of the so-called second wave that occurred in France during the fall of 2020.

## 2 Materials and methods

### 2.1 Data sources

#### 2.1.1 WWI

The local and regional values of WWI data are freely available for all plants treated by the Obepine network [here](https://www.data.gouv.fr/fr/datasets/surveillance-du-sars-cov-2-dans-les-eaux-usees-1/).

#### 2.1.2 Incidence rate

Incidence rate data are partially available in open access for 22 EPCIs and can be found [here](https://www.data.gouv.fr/fr/datasets/indicateurs-de-lactivite-epidemique-taux-dincidence-de-lepidemie-de-covid-19-par-metropole/). For the *Grand Reims* metropolitan area, incidence data are not available in open access. We have retrieved them by studying the different dashboards issued by the ARS Grand-Est ([example here](https://www.grand-est.ars.sante.fr/system/files/2021-04/2021-04-16_Données%20du%20jour.pdf)). For three additional plants (*Lagny-sur-Marne, Evry* and *Paris Seine Morée*), the data corresponding to the specific watershed of these plants were directly transmitted to us by *Santé Publique France*.

### 2.2 Data analysis

Statistical analyses were performed using R and Python programming languages. When not directly provided, the incidence rate was computed according to [the same formula](https://www.data.gouv.fr/fr/datasets/indicateurs-de-lactivite-epidemique-taux-dincidence-de-lepidemie-de-covid-19-par-metropole/) used by *Santé Publique France*, using a weekly moving average. Clinical data were then processed through [statsmodels’ seasonal decomposition function](https://www.statsmodels.org/stable/generated/statsmodels.tsa.seasonal.seasonal_decompose.html) to extract their trends. 24 WWTPs were considered in the different statistical analyses, with varying sampling frequency detailed later on.

### 2.3 Sampling, transport and analysis

The statistical studies of this document were carried on a part of our total French wastewater samples collected between March 3rd 2020 and May 1st 2021. The protocol is as the following: wastewater samples were taken integratedly during a 24-hour period, were conserved at 5°C (+/-3°C) and transported at 4°C. Quantification analyses, involving extraction, concentration and RT-qPCR or RT-dPCR steps [9, 10], were performed within 3 days after sampling. The data associated with these samples included incoming volume at the plant inlet, ammonium concentration, conductivity and COD^4^. The results of the quantification (in number of genome unit per liter) and other related data were then processed by mathematical tools. RT-qPCR or RT-dPCR were performed on the E and RdRp genes, the former being routinely used to process the WWI and the latter being used for validation purpose.

### 2.4 Consideration of flow fluctuations at the wastewater treatment plant inlet

The WWI can have different quality indices, or EDQPI,^5^ depending on the richness of the data provided. A quality index of 1 corresponds to a viral load level without taking into account the flow inlet of a WWTP. That of 2 is improved, compared to 1, by adjusting the WWI using the incoming volume information. This helps neutralise the dilution effects due to precipitation or to watershed deviation. A level equal to 3 suggests the use of other physicochemical factors like NH4^+^, conductivity and COD in order to induce the wastewater volume related to human activities. Detailed mathematical formulas are indicated later on.

The problem can be expressed as follows. Let *C*_0,*t*_ be the SARS-CoV-2 concentration in the water that arrives at the inlet of the treatment plant. Then the SARS-CoV-2 concentration without dilution effect impacting the nominal operation of the network can be computed as follows:

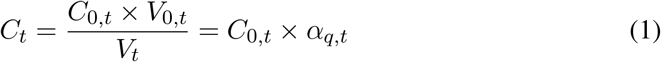

where *V*_0,*t*_ is the total volume at the inlet of the treatment plant on day *t* and *V*_*t*_ is the household wastewater volume. As these quantities need to be estimated, we approximate *α*_*q,t*_ by 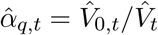, which is the volume normalization coefficient at time t and EDQPI q, where 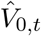 and 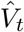 are the estimations used for *V*_0,*t*_ and *V*_*t*_, respectively.

- When *V*_0,*t*_ and *V*_*t*_ are both unknown, the EDQPI is, by design, equal to 1 and both volumes are approached by the mean daily incoming volume at the inlet of the WWTP. This volume *V*_*db*_ is extracted from the database of the French MTES^6^ listing all the useful data for the year 2017. We then have 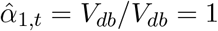.
- When *V*_0,*t*_ is measured and *V*_*t*_ is unknown, the EDQPI is, by design, equal to 2 and we approach *V*_*t*_ by *V*_*db*_. We then have 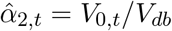.
- When *V*_0,*t*_ is measured and *V*_*t*_ is estimated from physico-chemical dilution indicators (such as NH4^+^ concentration, conductivity and COD), the EDQPI is, by design, equal to 3. We then have 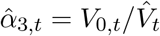.

When the EDQPI is equal to 3, *V*_*t*_ is estimated by the average between rectified volumes from ammonium, conductivity and COD :

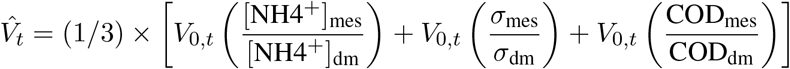

where *V*_0,*t*_ is the total volume at the inlet of the treatment plant on day *t*, [NH4^+^]_mes_ is the NH4^+^ concentration measured on day *t*, [NH4^+^]_dm_ is the mean concentration of NH4^+^ measured on dry conditions the previous year, *σ*_mes_ is the electric conductivity measured on day *t* in S.cm^−1^, *σ*_dm_ is the mean electric conductivity measured on dry conditions the previous year, COD_mes_ is the chemical oxygen demand measured on day *t*, COD_dm_ is the mean chemical oxygen demand measured on dry conditions the previous year.

This formula only applies to days when rainfall was recorded and no civil engineering of the wastewater network was involved. Indeed, these could have caused the daily incoming volume to be significantly weaker than the mean of the historical year used to assess physico-chemical concentrations in dry conditions, thus leading to an incorrect estimation of rainfall induced additional volume on rainy days.

In order to understand the importance of these additional data, we estimated by simulation the difference between the WWI with quality indices equal to 1 and 2. To do so, we first calibrated a parametrised statistic model under the two different settings of EDQPI 1 and 2, i.e., without and with inlet volume measurement respectively, hence we got two WWI curves of corresponding EDQPI. Then for each of the two statistic models, we simulated a group of 1000 trajectories from its parameters. We finally computed the root-mean-square (RMS) deviation between the WWI of EDQPI 2 and each curve of each group of simulated trajectories. With the two sets of RMS deviations, we performed a one-factor ANOVA test to assess the impact of absence of daily incoming volume measurement of a plant, with null hypothesis being no significant difference between the 2 groups. We conducted the study on 22 sewage plants each with several samples taken on rainy days with several months of history. We ran the same simulation to compare EDQPI 2 and 3, this time on 2 WWTPs for lack of sufficient physico-chemical data on the remaining sewage plants.

### 2.5 De-noising and interpolation through Kalman smoothing

RT-qPCR quantifications are subject to many uncertainties. Using only the calculated virus concentrations to monitor the pandemic can therefore be misleading, as a large increase in the measured concentration can be due either to a real increase in virus concentration or to a positive quantification error. This error can be caused by different factors, during the concentration, extraction or RT-qPCR phases, as well as during the integrated sampling at WWTP and its transportation. Thus, standard materials and laboratory practices have a strong influence on the RT-qPCR performance [2]. Moreover, the raw signal included in each person’s stool may be altered during its stay in the sewer system and during the aforementioned analysis steps [3]. This is why these data are pre-processed through Kalman smoothing [6, 7, 8] in order to provide an estimate of the real amount of virus and to evaluate the uncertainty on this estimate. In this method, the existence of a time dependency between the actual quantities is exploited (i.e. an actual virus quantity in the wastewater on a given day provides information about the quantity that will be observed on the following days, due to the outbreak dynamics), while the successive errors in virus concentration measurements are independent from each other.

The concentrations to be measured are sometimes below the quantification or the detection RT-qPCR thresholds. Consequently, we face a problem of censored data. In addition, samples are typically collected twice a week, resulting in missing data on some days. Finally, outliers may bias the smoothing. A new one dimensional Kalman smoothing method [4] has been developed to adapt to these particularities for the needs of Obepine, which implied a numerical discretization. We applied the developed smoother on the logarithm of the measured quantities in order to take into account the exponential character of the growth observed during the epidemic period and the heteroscedasticity observed empirically on the residuals when the method is applied directly.

The mathematical writing of the underlying model is as follows:

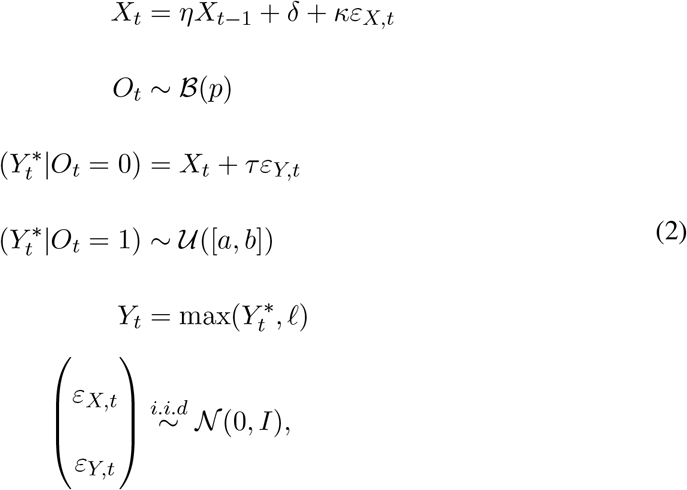

where:

*t* is the time index (ranging from 1 to *n* days), *X*_*t*_ ∈ ℝ is the logarithm of the real concentration in wastewater at time *t, X* = (*X*_*t*_)_*t*∈{1,…,*n*}_ is the vector of log-transformed real concentrations (to be recovered) and *Y*_*t*_ ∈ ℝ is the logarithm transformation of the estimated concentration in wastewater measured by RT-qPCR at time *t, C*_*t*_, defined in Equation 1 (*Y*_*t*_ = log(*C*_*t*_)). *Y*_*t*_ is generally only partially observed. We note 𝒯 ⊂ {1, …, *n*} the set of *t* at which *Y*_*t*_ is observed. *Y* = (*Y*_*t*_)_*t*∈𝒯_ is the vector of measurements. *Y* ^*^ is an accessory latent variable corresponding to a non-censored version of *Y*. *I* is the identity matrix. *η* ∈ ℝ, *δ* ∈ ℝ, *κ* ∈ ℝ^+^ and *τ* ∈ ℝ^+^ are parameters (to be estimated). *ℓ* is the threshold below which censorship applies^7^. *O*_*t*_ ∈ {0, 1} is, for any *t* ∈ 𝒯, the indicator variable of the event “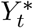 is an outlier”. *O* = (*O*_*t*_)_*t*∈𝒯_. ℬ(*p*) stands for the Bernouilli distribution of parameter *p* and 𝒰([*a, b*]) for the Uniform distribution on the interval [*a, b*]. *p* is a meta-parameter designating the a priori probability of being an outlier (we take *p* = 2% here). *a* and *b* have to be chosen, they can for example correspond to quantiles (respectively very close to 0 and very close to 1) of the empirical marginal distribution of *Y*. The parameters *η* ∈ ℝ, *δ* ∈ ℝ, *κ* ∈ ℝ^+^ and *τ* ∈ ℝ^+^ of maximum likelihood are estimated by numerical optimization through Nelder-Mead [5] as explained in [4]. At time *n*, the developed smoother gives the law of *X*_*t*_ for *t* ∈ {1, …, *n*} knowing *Y* = (*Y*_*t*_)_*t*∈𝒯_, as well as the probability for each *Y*_*t*_ to be an outlier. We note the produced reconstitution 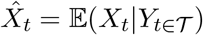.

### 2.6 Consideration of inter-laboratory variability

Several laboratories are providing sewage water SARS-CoV-2 viral load analyses to Obepine, each of them being in charge of various WWTPs. These laboratories have been selected based on their ability to carry out analyses properly using protocols that have been validated for the quantification of SARS-CoV-2 in wastewater [9, 10]. Nonetheless, comparative ILA^8^ have demonstrated that the estimated virus concentrations obtained on the same samples by different laboratories could sometimes differ in the order of magnitude of 1 log as shown in Table 2. In order to obtain a universal indicator for normalizing data provided by different laboratories [30], we have reworked the analysis results. The level of the indicator for a specific plant is thus related to the maximum concentration recorded by its associated lab on all the plants assigned to it within the Obepine network over a specific period. We have chosen a period between June 1st 2020 and January 1st 2021, which gives a maximum corresponding to the peak of the second wave of the epidemic. We then perform the following normalization:

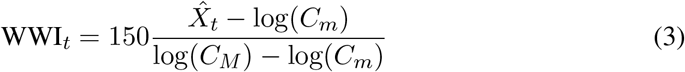

**Table 1:** Significance test results for difference between EDQPI 1 and EDQPI 2.

| WWTP | Inhabitant equivalent capacity | Number of samples per week | p-value |
| --- | --- | --- | --- |
| Forges-les-eaux | 16 000 | 1 | <0.0001 |
| Fécamp | 45 000 | 1 | <0.0001 |
| Saint-Denis lès Sens | 64 000 | 2 | <0.0001 |
| Auxerre-Appoigny | 83 000 | 2 | <0.0001 |
| Nantes-2-Petite Californie | 180 000 | 2 | <0.0001 |
| Evry | 220 000 | 2 | <0.0001 |
| Lyon-La Feyssine | 300 000 | 2 | <0.0001 |
| Le Havre | 320 000 | 1 | <0.0001 |
| Lagny-sur-Marne | 350 000 | 2 | 0.0017 |
| Dijon | 400 000 | 2 | <0.0001 |
| Lille Grimonpont | 420 000 | 2 | <0.0001 |
| Reims | 470 000 | 7 | <0.0001 |
| Nancy-Maxeville | 500 000 | 2 | <0.0001 |
| **Rouen** | 550 000 | 1 | **0.488** |
| Paris Marne Aval | 550 000 | 2 | <0.0001 |
| Nantes-1-Tougas | 600 000 | 2 | <0.0001 |
| Nice-Haliotis | 620 000 | 2 | <0.0001 |
| Lyon-Pierre Bénite | 630 000 | 2 | <0.0001 |
| Toulouse-Ginestous | 950 000 | 2 | 0.034 |
| Lyon-Saint-Fons | 980 000 | 2 | <0.0001 |
| Strasbourg | 1 000 000 | 2 | 0.0012 |
| Paris Seine Amont | 3 600 000 | 2 | <0.0001 |

**Table 2:** May 2021 ILA results as scaling factors between the 9 Obepine laboratories, in relation to one laboratory taken as reference (Lab 1).

|  | Lab 2 | Lab 3 | Lab 4 | Lab 5 | Lab 6 | Lab 7 | Lab 8 | Lab 9 |
| --- | --- | --- | --- | --- | --- | --- | --- | --- |
| Scaling factor mean | 0.95 | 1.20 | 1.96 | 3.96 | 10.64 | 0.40 | 6.50 | 1.12 |
| Scaling factor std | 0.31 | 0.34 | 0.54 | 0.74 | 9.00 | 0.077 | 2.62 | 0.43 |

Where WWI_*t*_ is the WWI value at time *t*, 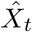 is the previously defined reconstitution, *C*_*m*_ represents a quantification threshold of 1000 GU/L and *C*_*M*_ is the maximum concentration historically recorded by the reference laboratory on plants with average daily flows similar to that of the plant of interest. The normalization factor of 150 was chosen a posteriori, so as to obtain a level between 40 and 85 around the beginning of September 2020, a period which corresponds for the majority of the plants to the middle of the exponential growth phase of the second wave in France. This level corresponds to a circulation level between fairly low and average, which would have given enough time to alert on the situation of resumption of the epidemic at this time. The maximum concentration is not solely based on the laboratory’s history, but more specifically on the basis of plants with a similar flow to the one to be standardized. This additional selection makes it possible to harden the comparison criterion and to strengthen the ability to compare agglomerations where the epidemic situation is similar. For example, it is more likely to have 80% of the population infected at the same time in a sewage plant treating 10 inhabitants than in a sewage plant treating 10 million people. Without this partitioning, there could be a problem of underestimation of the epidemic situation in very large agglomerations in case of a critical health situation at a WWTP of much more moderate size, since the maximum concentration could never be approached by large sewage plants. We then chose to split the sewage plants in ten bins according to their average daily incoming volume, and assign a maximum concentration to each category.

This formula still had a major drawback in the case of laboratories joining the project later than the historical ones, typically after December 2020. To deal with this flaw, we ran several ILA which we used to assess and update a proportionality coefficient between laboratories running the same protocol. For a laboratory joining late with no historical record, we multiply its analysis results by this proportionality coefficient and use the *C*_*M*_ of the laboratory we have chosen as the reference for the calculation of this coefficient. Finally, under logistics and transport constraints and the workload limit of the laboratories, we designed that each laboratory receives and analyses sewage samples from plants distributed as evenly as possible over the French territory. This choice avoids the situation where one laboratory is assigned only to cities with a low incidence of the disease and another to cities with a high incidence of the disease, a situation that would make difficult to compare the level of virus circulation between them. The consideration of this inter-laboratory variability allowed us to aggregate the WWI of different WWTPs and elaborate regional indicators to have a more objective insight of the epidemic situation on a larger scale. Each regional indicator represents the weighted average of the local indicators in the same area, with the weight of each plant corresponding to its average daily volume.

## 3 Results

We propose herein a new indicator (WWI) to convert the estimated amount of viral genomes that enter a WWTP per day in a unitless value. Diverse mathematical models (see Materials and Methods) make it possible to propose a smoothed tendency curve that faithfully reflects the epidemic situation at a WWTP.

### 3.1 De-noising and interpolation through Kalman smoothing

The results of this pre-processing are illustrated on an example of simulated data on Figure 2 and on an example of real data from the Obepine network on Figure 3. As shown in Figure 2 on a set of simulated data, the mean signal reconstituted through this model faithfully reflects the true underlying process and shows low sensitivity to outliers. The successive reconstitutions of the underlying “true” auto-regressive process are expected to change at each new data point, since those bring additional information with regard to the past. This is depicted Figure 3, with successive reconstitutions in different colors. Each intermediary reconstitution lies inside the 95% prediction interval of the final reconstitution. The difference between the final reconstitution and each of the intermediary reconstitutions is quite low, which means that there is usually not a lot of difference between the results transmitted at a given date and those transmitted a week later with a pair of additional data points.

**Figure 2:**
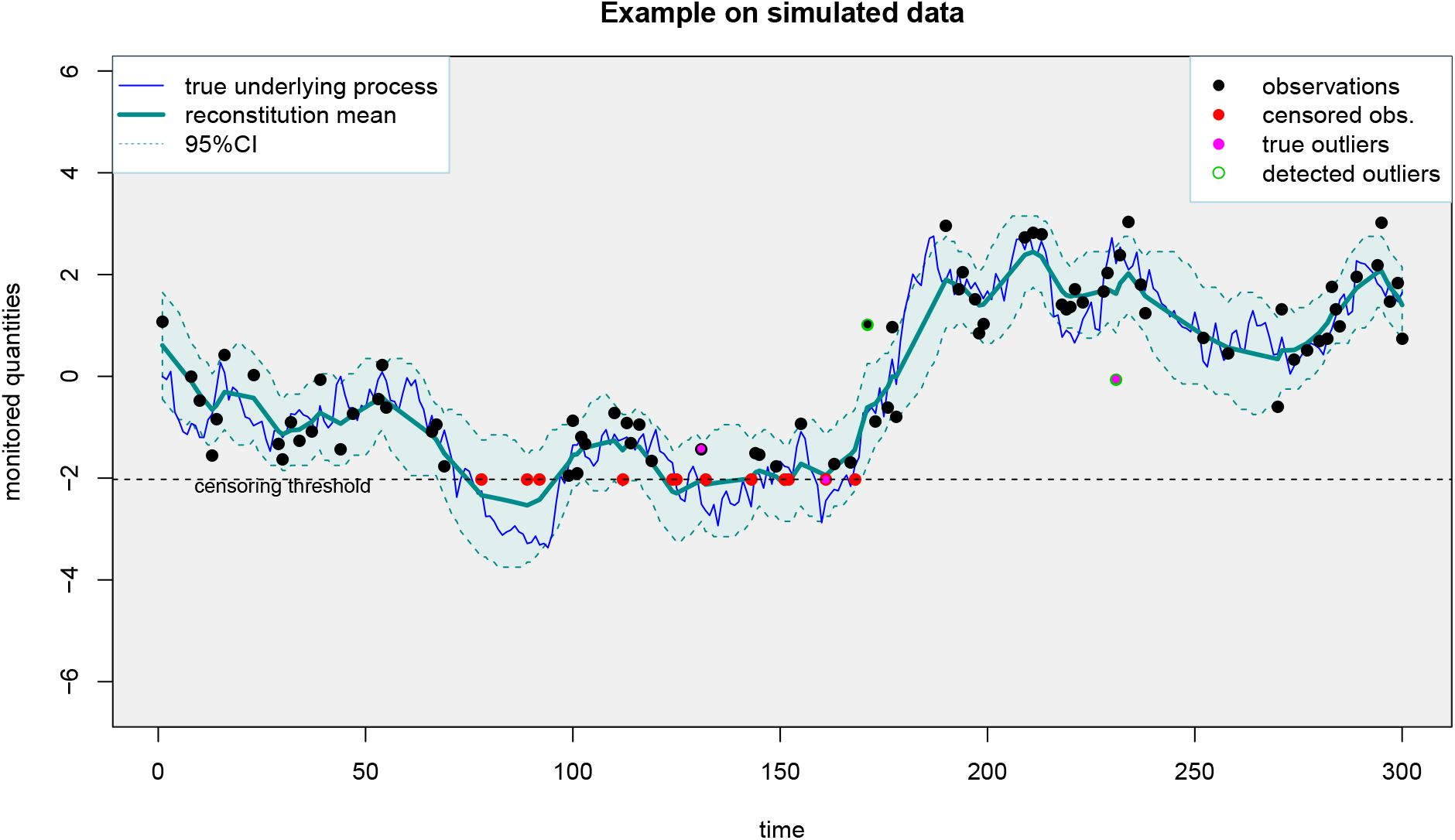
An example of the application of the proposed smoother (taking into account censoring and outliers) on simulated data with 16% of censored data and *p* = 2% of outliers. The censoring threshold corresponds to the RT-qPCR quantification threshold. The 95% prediction interval should cover about 95% of the true underlying process (blue curve). The mean reconstitution is faithful to the true underlying process.

**Figure 3:**
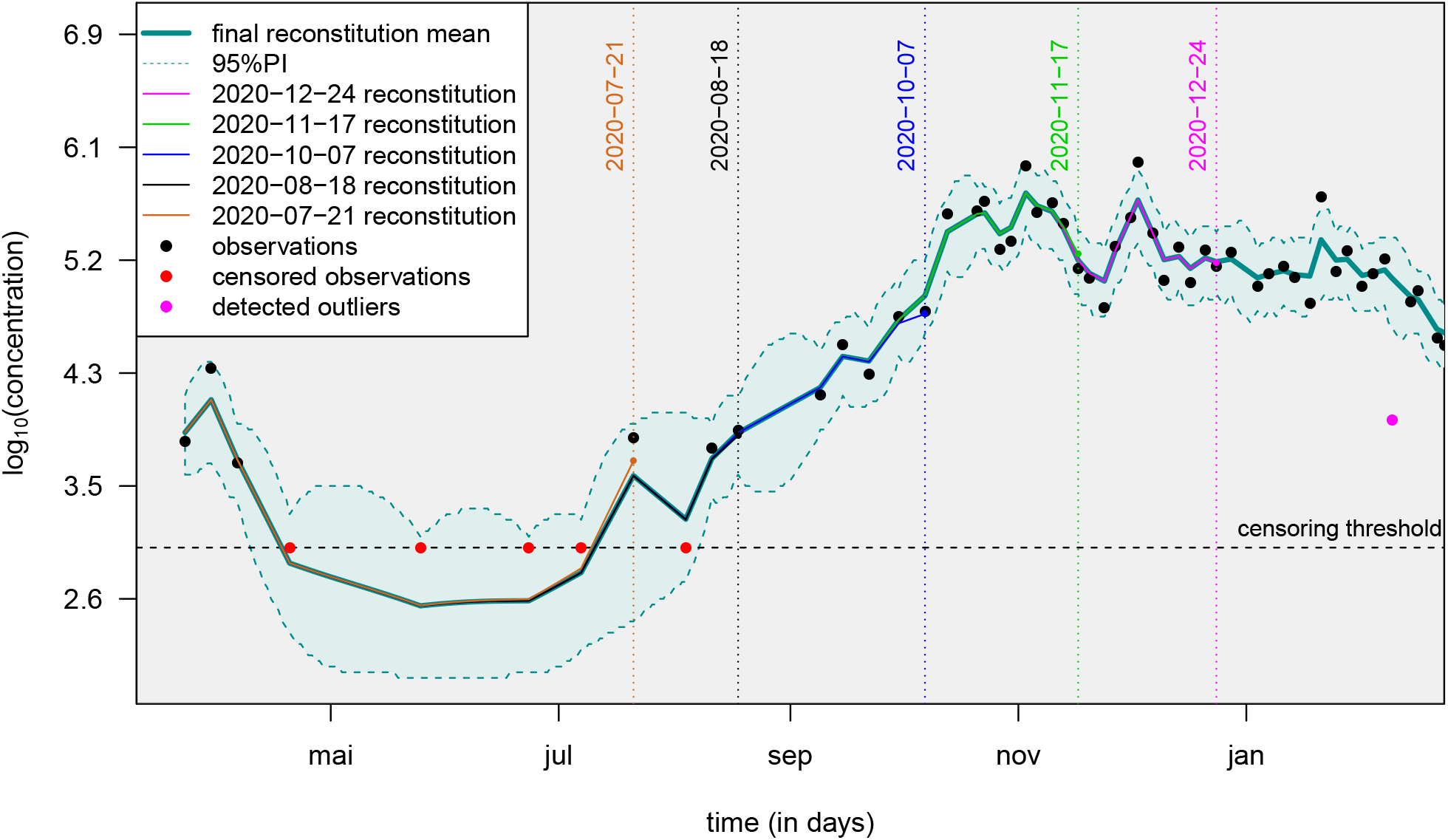
An example of the application of the proposed smoother (taking into account censoring and outliers) on data from a wastewater treatment plant of the Obepine network : successive predictions for the underlying process (never observed), *X*, 95% prediction interval and detected outliers (with an outlier proportion of *p* = 2%). The censoring threshold corresponds to the RT-qPCR quantification threshold. Each vertical dotted line corresponds to intermediary reconstitutions over the course of the project, without taking into account any additional data point past the reconstitution date. The difference between these intermediary reconstitutions and the final reconstitution gives an idea of the error made weekly prior to knowing future data points. The WWTP is the one in charge of the EPCI of *Dijon*, its associated laboratory being Lab 2, see Table 2.

### 3.2 Impact of inflow variation on the WWI

Each trend curve is associated with a reliability index (EDQPI). EDQPI equals 1 when the WWI is calculated with an estimated flow and 2 when the real wastewater flow is used. By using the actual inflow volume of a plant, dilution effects by one-time events such as precipitation and civil engineering on the sewage network can be counterweighted. This led us to estimate the impact of rainfall on local trend curves. Table 1 shows that the difference between WWI signals calculated with EDQPI 1 and EDQPI 2 data is statistically significant in 21 of 22 WWTPs. The only case for which the null hypothesis is not rejected is *Rouen*, which is one of the plants sampled only once a week. With an average of 180 rainy days per year, it is conceivable that the test result would be different with a higher sampling frequency. Therefore, this result indicates that plant inflows needs to be informed as soon as possible to improve EDQPI and primarily during periods of prolonged rainfall or reduced flow, regardless of plant size. We also tested the differences between quality indices 2 and 3 at two plants. EDQPI is set to 3 when physico-chemical factors can be measured on samples such as NH4^+^ concentration, conductivity and COD. The ANOVA results suggest that the difference is not significant this time (i.e., an EDQPI of 2 would be as effective in accounting for rainfall as an EDQPI of 3) although further investigation on a larger number and a wider variety of plants would be required.

### 3.3 Consideration of inter-laboratory variability

We take a critical look at the normalization technique we used to account for the inter-laboratory variability. As no WWTP had been analyzed by at least two different laboratories over the course of the project, we simulated an hypothetic behavior of a network with only one plant analyzed by the 9 Obepine laboratories. We chose one laboratory as a reference (Lab 1), and simulated quantification results varying from this reference, using May 2021 ILA results summarized in Table 2. To do so, we simulated a synthetic signal and assigned it to Lab 1. Then, using Table 2, we synthesized 8 others signals using scaling factors drawn from normal distributions whose parameters were estimated using May 2021 ILA results. For each sampling date and each laboratory, a credible scaling factor was drawn from these normal distributions. We compared three normalization techniques. CM refers to a single common maximum concentration among all laboratories. LSM refers to the modelisation we used, with a laboratory-specific maximum concentration. CMILA refers to a single common maximum concentration after scaling all the laboratories results to a reference laboratory using ILA results. Figure 4 shows that our normalization technique significantly reduces the inter-laboratory variability for laboratories 4 to 8. Results are not significantly improved for the remaining 3 laboratories because such a normalization is not needed, as their scaling factors are close to 1 and their inter-samples replicability is quite good. Results can still be significantly improved, especially for lower values of WWI, once ILA are carried out.

**Figure 4:**
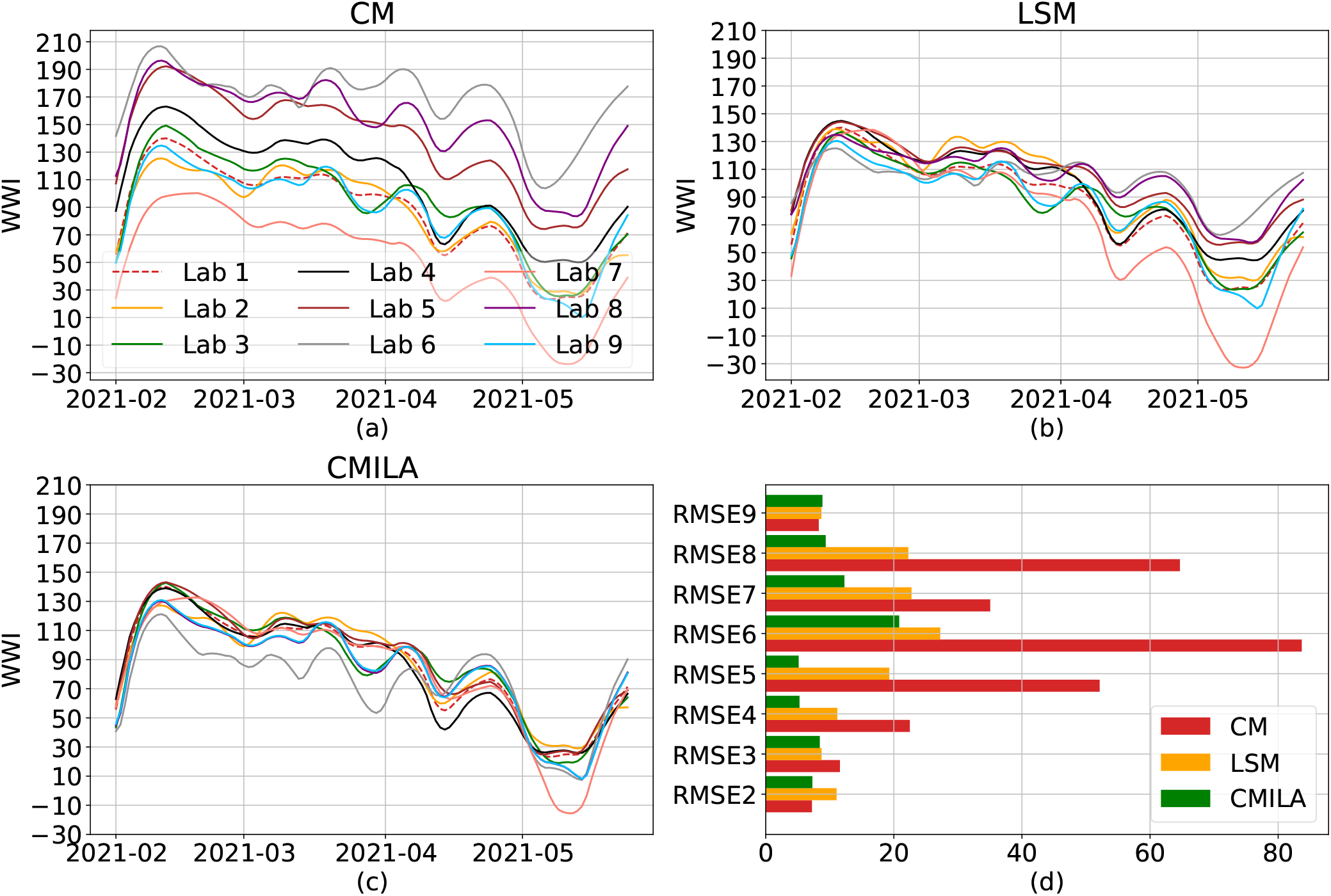
Simulation of different inter-laboratory variabilities and normalization techniques. We simulate the simple case of a single plant in a network analyzed by the 9 Obépine laboratories. (a) shows the results if the WWI normalization formula is applied with a *C*_*M*_ common to all laboratories. Results show a clear disparity between laboratories and a strong attenuation towards laboratories with lower quantifications results than laboratory 6. (b) shows the correction brought by using a *C*_*M*_ specific to each laboratory. Results are significantly improved for laboratories 4 to 8. The difference is not significant for the remaining 3 laboratories which all have a scaling factor close to 1 and a good inter-samples replicability. (c) shows the correction brought by using ILA results and estimating a scaling factor between each laboratory and Lab 1. As shown in (d), CMILA still is the overall best normalization technique. CM, LSM and CMILA respectively accounts for a common maximum, a laboratory-specific maximum and a common maximum after scaling following ILA. RMSE are calculated using the Lab 1 as reference.

### 3.4 Correlation and lag between the WWI and the incidence rate

We now focus on several EPCIs whose incidence rate is available at a local scale, within which the sewage network connects to one single WWTP, to limit possible omission biases related to the outbreak of the epidemic in neighborhoods not connected to the monitored plant, which may produce a phase shift. To determine the period over which to calculate the correlation between the two signals (the WWI and the incidence rate), we consider the following. The results of the virological tests are reported by municipality of residence and not municipality of testing, while the wastewater signal is localized and unchangeable. Moreover, the WWI is expected to capture contributions from asymptomatic and mildly symptomatic patients, which we suspect not to be negligible during the June-August 2020 period, whereas the incidence rate only reports diagnosed people. As we want to calculate the correlation between the two signals over a period where they are supposed to be similar and thus where the WWI is supposed to mainly capture a majority of people also likely to be diagnosed, we decided to focus on the period corresponding to the second wave of the epidemic in France. To avoid being biased by the movements of individuals during the 2020 summer vacations, we consider the start date of September the 1st, 2020, from which the majority of holidaymakers returned to their residence city. We consider that the last point of the interval of interest is the date from which the signal undergoes a new growth phase following the decay of the second peak of the epidemic. This date can thus vary depending on the different local dynamics of the epidemic. We then drag the subpart of the incidence rate curve over a +/- 30-day window until we find the time lag that yields the best correlation with the WWI. We use cross-correlation as a measure of similarity between the two signals. The cross-correlation calculation is performed between the WWI and the log transformation of the incidence rate. Since correlation is sensitive to outliers especially when sample size is small, we subsampled the incidence signal using 50% of the available data so as to avoid certain special patterns resulting in an unnaturally high correlation. The time lag resulting the highest positive correlation is recorded. A positive lag value indicates that the WWI is ahead of the studied epidemic signal. A negative lag value indicates that the WWI is lagging behind it. We selected several EPCIs to study the results on cities of different sizes and various regions, using the results of three different laboratories. Finally, we briefly discuss the case of two regional WWIs.

Figure 5 shows an example of simulation results on the *Lagny-sur-Marne* WWTP. There is a strong correlation (> 0.92) between the WWI and the incidence rate during the second wave for this WWTP. Moreover, the optimal phase shift between the two signals is quite low (2 days), meaning the WWI was a great surrogate to the incidence rate at that time. Figure 6 and Table 3 show some interplant variance on the time lag and the correlation between WWI and incidence rate. Such a variance in time lag between WWTPs has already been reported [20]. The intra-experimental variance is significantly higher for the WWTP of *Nancy*, whose average correlation with the incidence rate is not as strong as that of the other WWTPs. As the samples were taken with a one shot sampling and not integrated over 24 hours until October 20th, 2020 at this plant, it cannot be excluded that the correlation is weaker due to a more pronounced noise on the samples taken before this date [12]. As previously argued, we did not consider the time period between July and August 2020, one of the reasons is that we may have detected an earlier emergence of the pandemic than the incidence rate, as witnessed before by [22]. An explanation could be that, by the time, it was mainly younger populations that were affected, among which less symptomatic cases were reported. It is then sensible that the proportion of tested positive to total infected was rather low at that time. It is thus conceivable that the signal captured by the WWI differs more significantly from the incidence rate during that period because the two indicators monitored different populations by that time than at the second peak of the epidemic. Such a change in the demographic of the pandemic has already been reported in the state of Massachusetts [19] and is shown in Figure 7. The correlation is still good between the two compared signals (>0.85 for every WWTP except Nancy), which is consistent with the results of [1, 9, 12, 25, 26, 27, 28]. An inter-WWTP variance in median time lag remains, as seen in Table 3, and is going to be discussed in section 3.5. Yet imperfect as they do not sample a population as large as the one surveyed by the incidence rate because we could only monitor a fraction of the cities of the different French regions, regional wastewater indicators still show a good correlation (minimum correlation of 0.8) with their clinical counterparts, as shown in Figure 8 and Table 4. Moreover, the regional WWI is peaking ahead of the regional hospitalizations for both studied regions during the second wave, which is consistent with the findings of [23, 24].This illustrates the good aggregation capability of the WWI thanks to the normalization techniques we used, and our ability to follow the epidemic situation at a larger scale, despite monitoring at best less than 60% of a region’s inhabitants, as shown in Table 4.

**Table 3:**
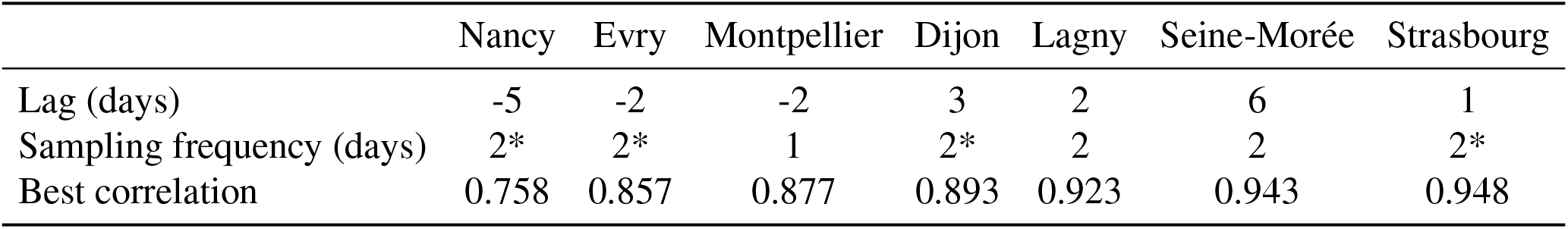
WWI and incidence rate lag estimates during the second wave of Fall 2020. Best correlation is the median of the best correlation over 1000 experiments. *Montpellier* was sampled once a week at that time. \**Strasbourg, Nancy, Evry* and *Dijon* were sampled once a week until mid October 2020, then twice a week. *Lagny* and *Seine-Morée* were sampled twice a week.

**Table 4:** Regional WWI correlation and lag estimates with incidence rate and hospitalizations during the second wave of Fall 2020. Best correlation is the median of the best correlation over 1000 experiments. IR means the WWI is compared with the incidence rate, H means the WWI is compared with the daily new hospitalizations in the corresponding region. The estimated surveyed population was calculated by considering the volume *V*_*db*_ of each plant and a daily consumption of 200L per inhabitant.

|  | Île-de-France - IR | Île-de-France - H | Grand-Est - IR | Grand-Est - H |
| --- | --- | --- | --- | --- |
| Lag (days) | -2 | 7 | 2 | 8 |
| Estimated surveyed population | 33.1% | 33.1% | 58.6% | 58.6% |
| Best correlation | 0.806 | 0.855 | 0.941 | 0.966 |

**Figure 5:**
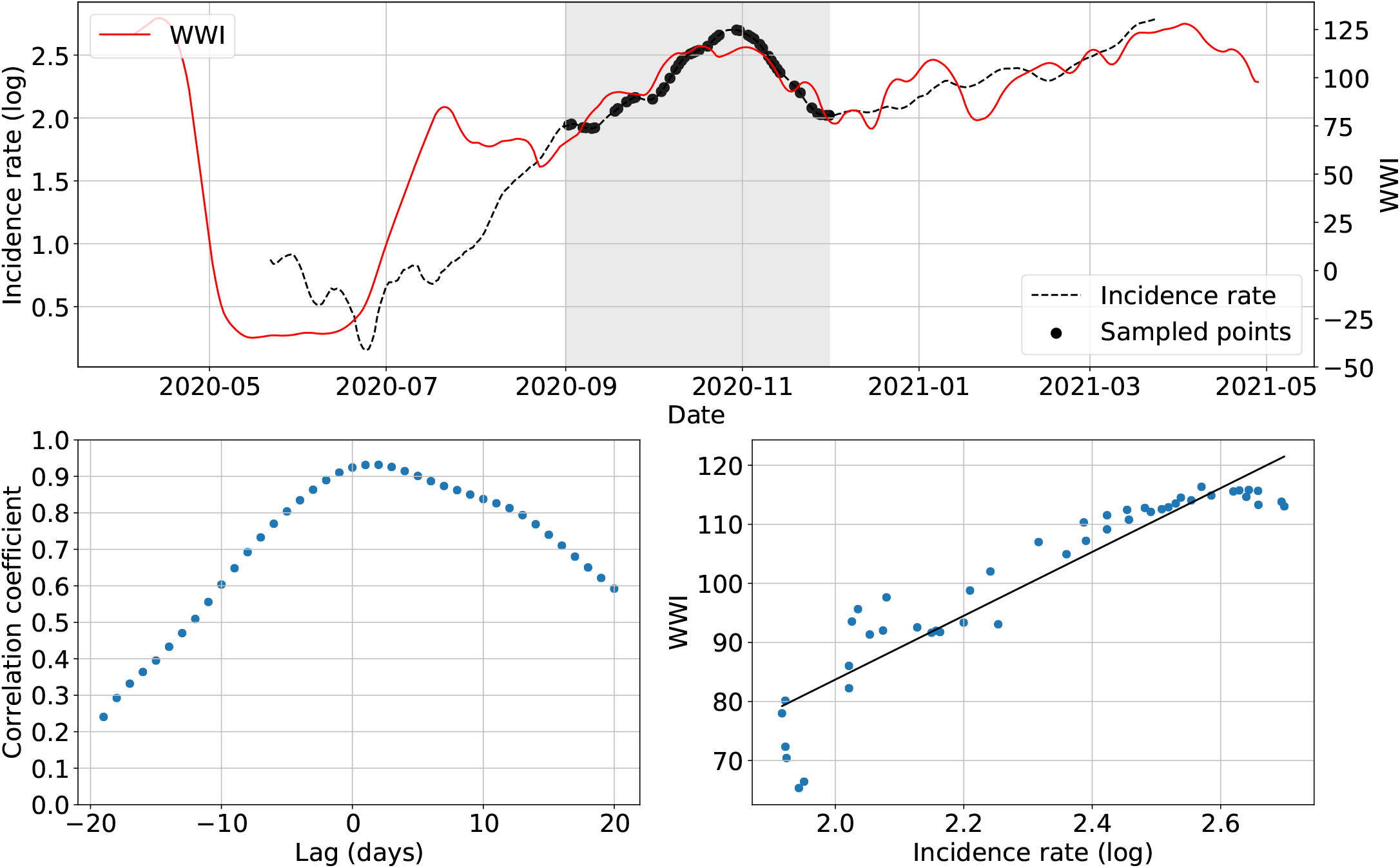
Simulation example on the *Lagny-sur-Marne* WWTP. The top plot shows WWI and incidence rate curves as well as the sample points selected for that simulation (the shadowy area corresponds to the period of interest). The bottom left plot displays the computed correlation values for lag values varying between -20 and 20 days. A positive lag means that the WWI is ahead of the incidence rate. A negative lag means that the WWI is lagging behind the incidence rate. The bottom right plot displays a scatter plot of WWI vs incidence rate at best time lag (2 days, with a correlation coefficient of 0.932), as well as the linear regression fitted on the data.

**Figure 6:**
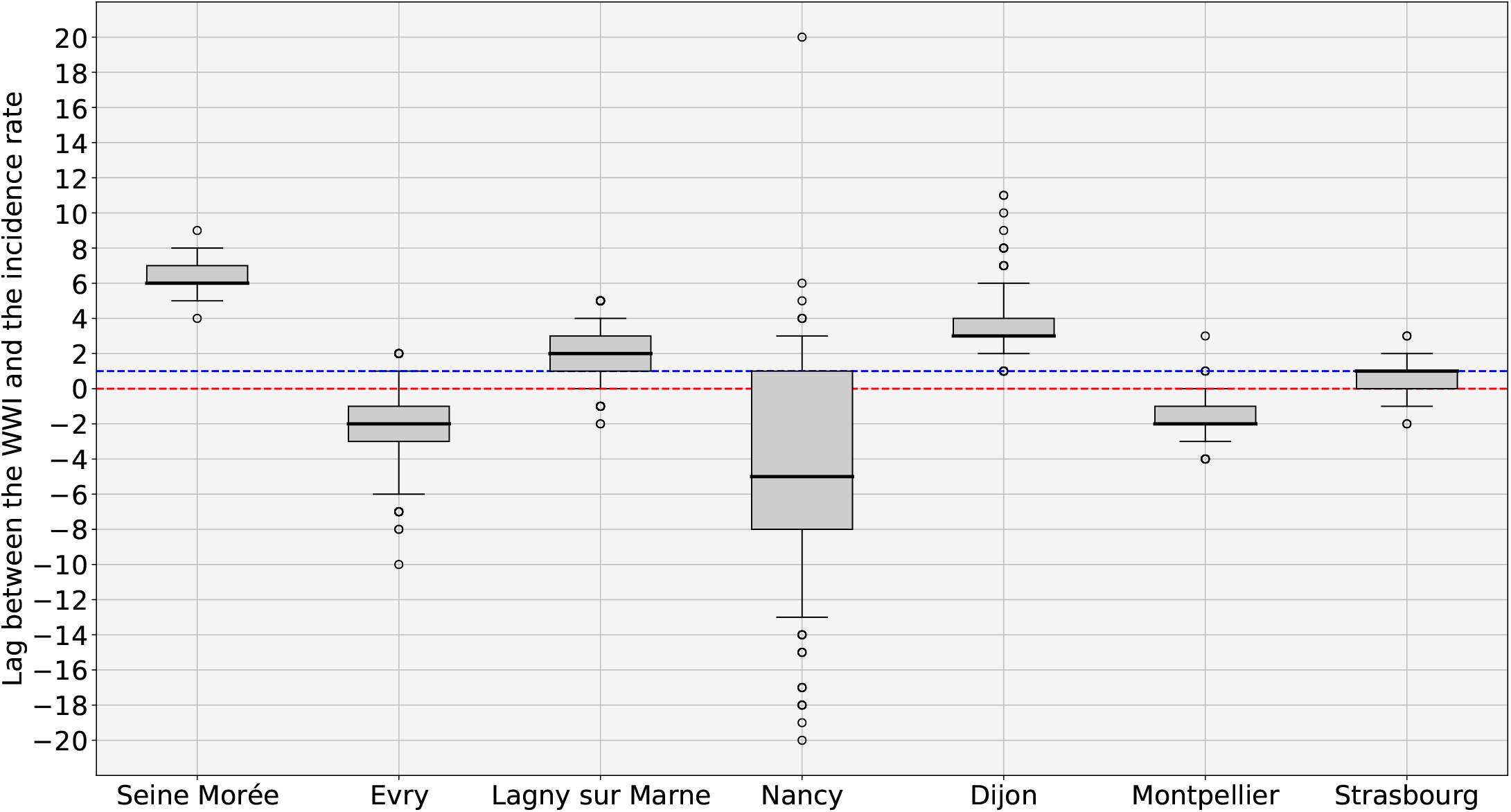
WWI and incidence rate lag estimates in days (*n* = 1000 simulations with random sampling of 50% of incidence rate curve). The Red dotted line indicates the zero offset level. The Blue dotted line is the median level over the 7 medians. The intra-experimental variance is significantly higher for the WWTP of *Nancy*, whose samples were not integrated before October 20th 2020, leading to a more pronounced noise on the first half of the wave.

**Figure 7:**
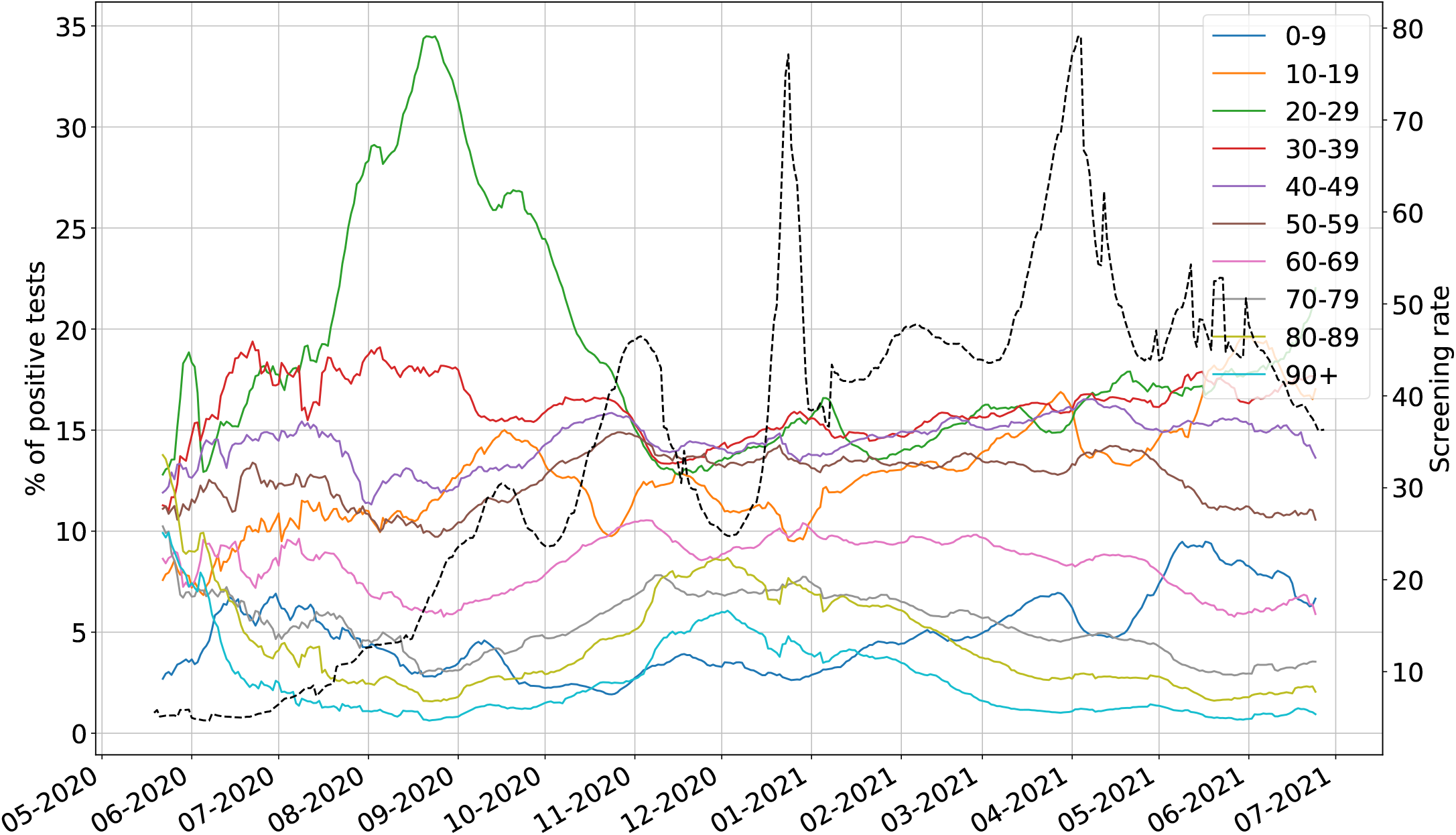
Evolution of the ratio of positive tests among each age bracket in France (straight lines) and of the screening rate (black dotted line). The screening rate corresponds to the number of test performed in France per 100,000 inhabitants. 20-29 years old bracket peaked during Summer 2020 and accounted for around 35% of the positive tests at its peak on August 21st 2020. Overall, the ratio increased from early June 2020 to late August 2020 among this age bracket. Conversely, the ratios among 40 years old and older categories were dwindling from July or even earlier for some of them. Infections were thus predominant among young people during Summer 2020 and less likely to be detected through conventional testing as the screening rate was about 3 times less important than at the peak of the second wave.

**Figure 8:**
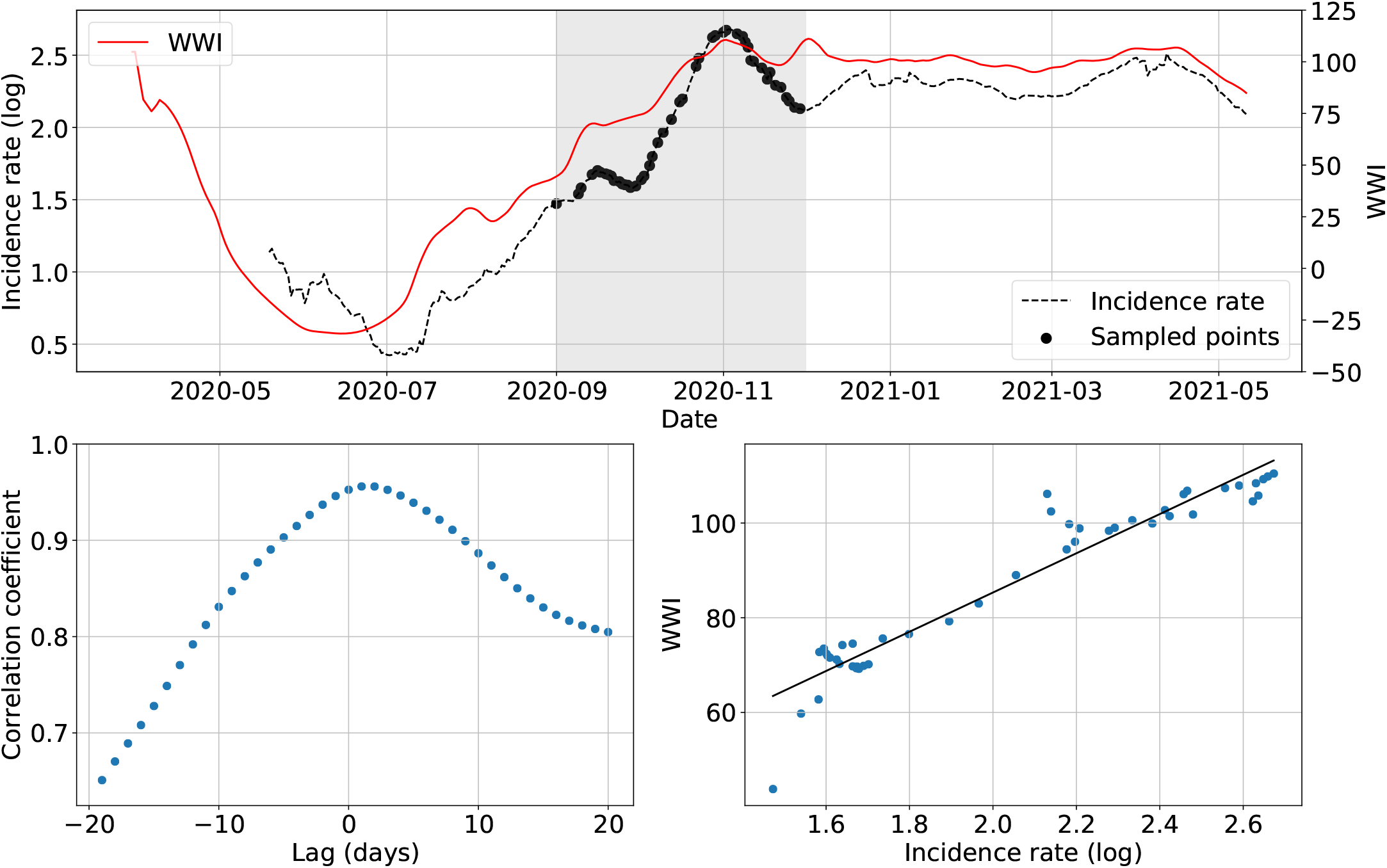
Simulation example for the Grand-Est region and the incidence rate. The top plot shows WWI and incidence rate curves as well as the sample points selected for that simulation (the shadowy area corresponds to the period of interest). The bottom left plot displays the computed correlation values for lag values varying between -20 and 20 days. A positive lag means that the WWI is ahead of the incidence rate. A negative lag means that the WWI is lagging behind the incidence rate. The bottom right plot displays a scatter plot of WWI vs incidence rate at best time lag (1 day, with a correlation coefficient of 0.956), as well as the linear regression fitted on the data.

### 3.5 Impact of the sampling frequency

The monitored WWTPs are collected twice a week with integrated 24h sampling, except for a few rare exceptions including the *Reims* WWTP, which is analyzed on average every day of the week, with rare exceptions. Since the *Reims* WWTP has been monitored for more than a year, it can be used to study the impact of the sampling frequency on the WWI signal. To do so, we compared its WWI signal with all available samples to WWI signals that would have been obtained from different sampling combinations comprised between 1 and 6 days per week. For the two-day tests, we only considered the case where the selected days were not consecutive. For the three-day simulation, we also prevented combinations where two days were consecutive. For the four-day scenario, we considered all possibilities except those where at least three days were consecutive. We then used two metrics to quantify this impact: RMSE between each WWI signal and cover rate between their respective 95% prediction intervals. We define the cover rate CR with the following formula :

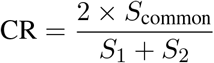

where *S*_common_ is the intersection area between the two prediction intervals (see Figure 10), *S*_1_ and *S*_2_ being the areas of the prediction intervals of the considered models. We chose this formula and not only the *S*_common_ to account for the case where wider prediction intervals, implying greater uncertainties, would lead to greater cover rates than better models with narrower intervals because it would have a greater intersection with the whole prediction interval of the default model.

**Figure 9:**
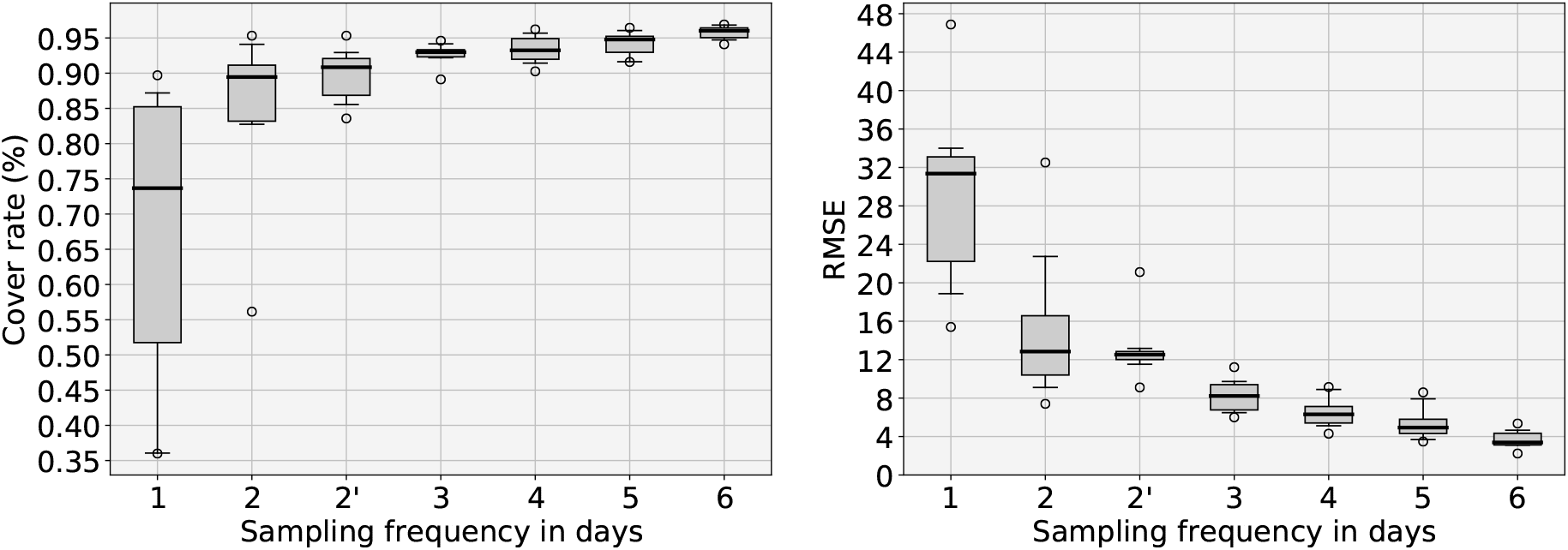
Quantitative results of the sampling frequency analysis performed over the *Reims* WWTP. The left plot displays the evolution of the cover rate between 95% prediction intervals obtained with a reduced number of sampling days and the full signal. The cover rate represents the common surface of 95% prediction intervals between the default model and the studied subsampled model. The right plot shows the RMSE between the WWI. The x-axis represents the sampling frequency. 2’ frequency is a particular case of biweekly sampling where at least 2 days separate each sampling day (e.g. Monday can only be paired with Thursday or Friday). 3 days sampling seems to be the best cost-performance tradeoff. 2’ solution still brings an improvement to simple 2 days sampling if 3 days sampling cannot be achieved.

**Figure 10:**
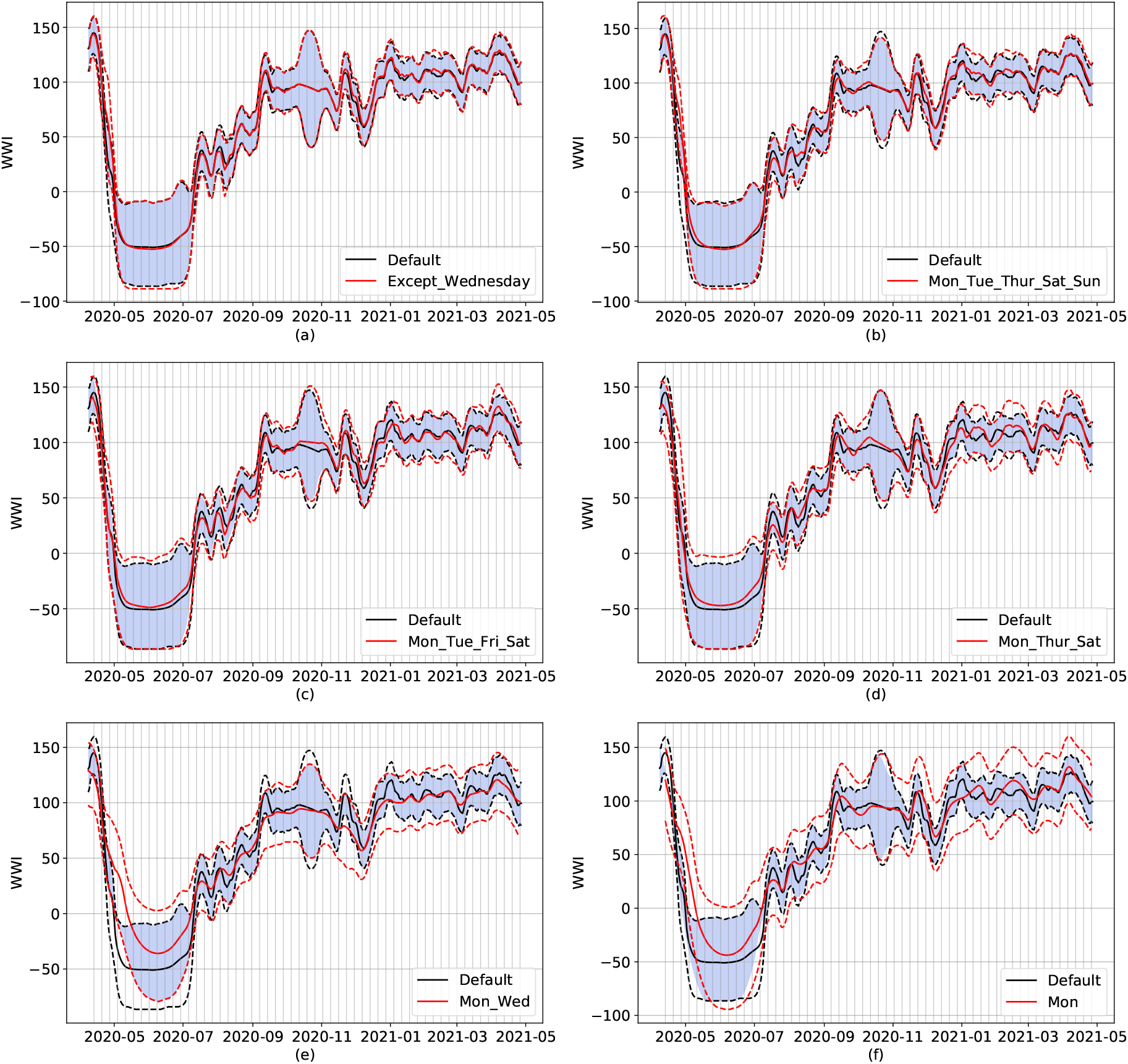
Examples of subsampling on the *Reims* WWTP, ranging from six days (top left) to one day per week (bottom right). Dotted lines represent the respective 95% prediction intervals for default (black) and subsampled (red) models. The default model uses all the available data from the *Reims* WWTP (usually 7 samples a week). Continuous lines show the WWI of both models. The blue-colored surface represents the intersection of both prediction intervals. The vertical grid corresponds to Mondays. On figure (d), short term trend of red and black signals differs early January. On subfigure (e), local peaks on early September and early December are missing on the subsampled signal. Subsampling can also induce couple days of time lags in peaks, as shown in figure (f) with both same local peaks.

Since the medians of the lags between the WWI and the incidence rate were quite different between WWTPs as shown in Figure 6, we wanted to evaluate the impact of the sampling days on this offset. To do so, we also used the data from the *Reims* WWTP. This allowed us to compare different versions of the WWI and to compare them with the incidence rate. We tested all combinations of two sampling days per week, excluding the possibility that sampling occurs on two consecutive days (a situation that can sometimes occur for logistical reasons but should remain exceptional). This plant was not included in the second wave offset study because wastewater analysis results were impacted by logistical problems at that moment. To assess the influence of sampling, we tested the time period around the January 2021 epidemic growth (between November 30th 2020 and January 22nd 2021), which is visible on both the incidence rate curve and the WWI curve. As the incidence data from *Reims* were not available for weekends and holidays, we revised the sampling rate upwards for the tests in this city as the number of points was lower (60% of the points compared to 50% for the studies focused on the second wave).

We can see on Figure 9 that both metrics show a clear improvement between once and twice a week sampling (RMSE is cut by more than half and median cover rate improves by 16%). While both RMSE and cover rate gains seem to be weaker than the ones we had from once to twice a week, it is important to notice that their variance has also been significantly reduced when upgrading from twice to three times a week. Achieved gains from three days and a more important sampling frequency does not seem as much interesting, for both metrics.

Qualitative wise, we can see on Figure 10 that going from 6 to 3 sampling days does not bring any significant difference to the WWI signal. Yet, short term interpretations can still be affected on specific periods as, the less sampling days available, the more biased towards outliers the WWI can become. Such a situation can be seen on subfigure (d): while the default signal is continuously dwindling from early to mid-January, the subsampled signal is actually shortly going down then increasing towards a plateau. Even though the general dynamics of the signal are still captured through once and twice a week sampling, local variations can be missed. On subfigure (e), local peaks on early September and late November are missing on the subsampled signal. They are captured through once a week sampling, but with a slight offset.

Figure 11 shows that a similar variance as the inter-WWTP variance shown in Figure 6 can be observed by changing the sampling days of the same sewage plant (the experiments were conducted on the *Reims* WWTP). Indeed, the difference in variance between the two sets of median time lags from the 7 WWTPs of Figure 6 and the 14 two-days combinations of Figure 11 is not statistically significant (p-value=0.78). The difference in time lags observed in Figure 6 between the 7 WWTPs studied could thus be notably explained by the approximation on the WWI signal because of subsampling.

**Figure 11:**
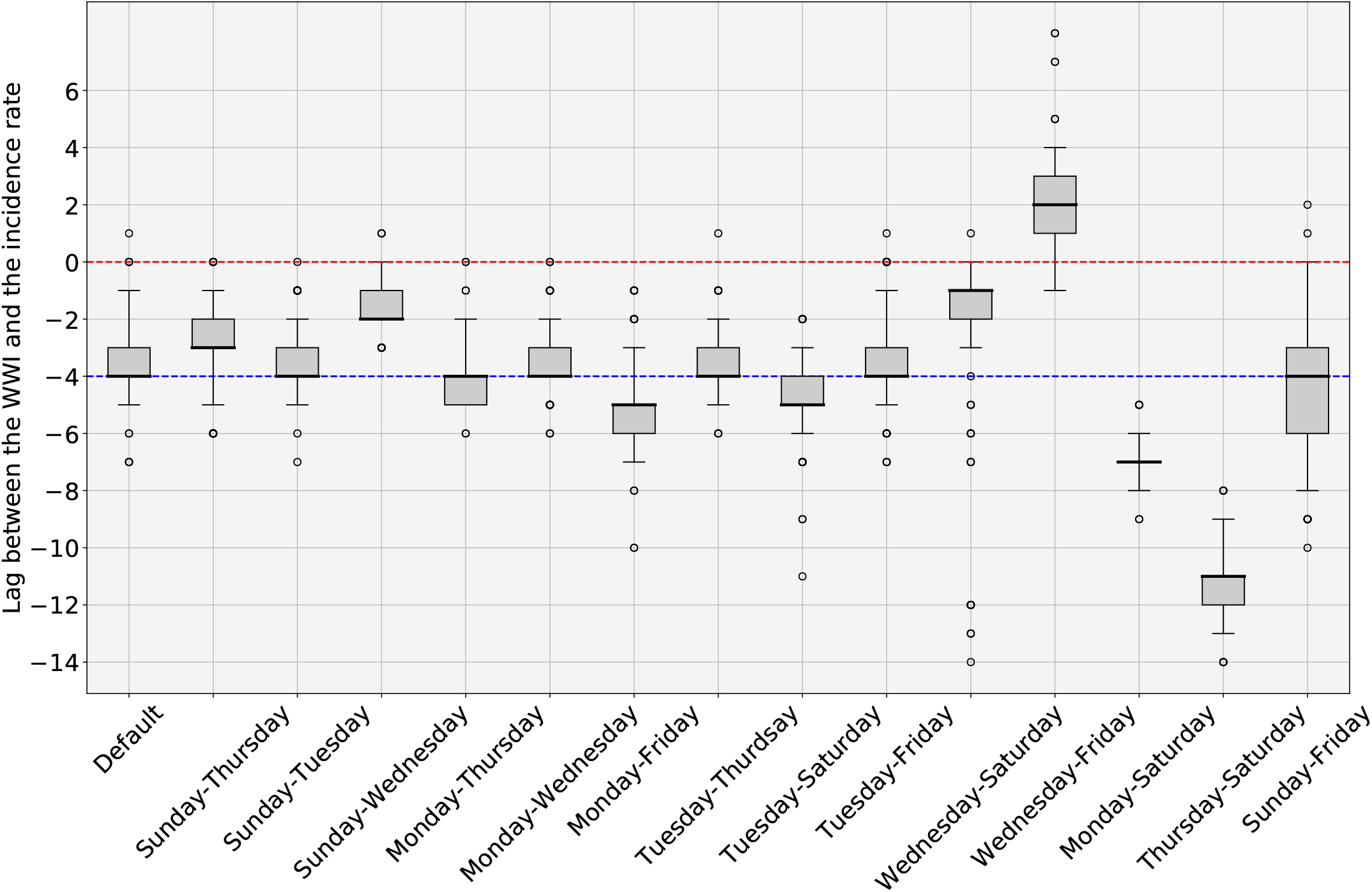
WWI and incidence rate lag estimates in days with varying sample days for the treatment plant of *Reims* (*n* = 1000 simulations with random sampling of 60% of incidence rate curve). Default corresponds to the WWI as it is routinely processed with every single data point available. Other possibilities are obtained through resampling twice a week on specific weekdays. The Red dotted line indicates the zero offset level. The Blue dotted line is the median level over the 14 medians. As the difference in variance between the set of median time lags from the 7 WWTPs of Figure 6 and the set of median time lags from the 14 two-days combinations displayed here is not statistically significant, subsampling could be one of the factors explaining the variability in optimal time lags between WWTPs shown in Figure 6.

**Figure 12:**
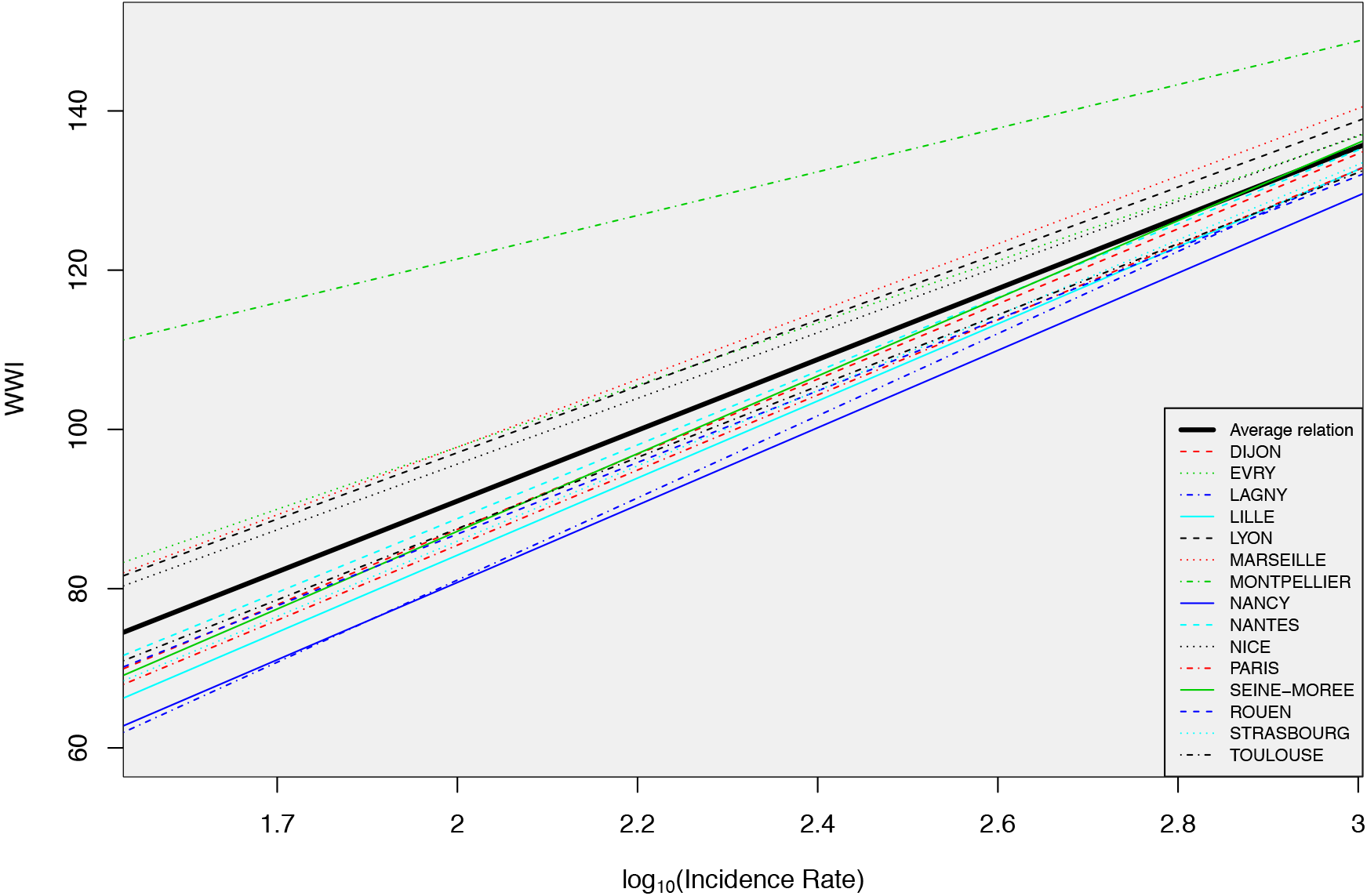
Relation between the WWI and the incidence rate in log scale learned by the full mixed effects model (Model 2). *Montpellier* relation greatly deviates from the average one. The significant deviation in intercept for *Montpellier* is probably due to an insufficient coverage of the French territory by the relative laboratory of this WWTP. The WWTP of *Paris Seine-Amont* was used for the comparison with the *Grand Paris* incidence rate.

### 3.6 Assessment of the comparative ability of the WWI

The WWI was designed to make comparable the analysis results provided by different laboratories, each with its own analysis bias. These plants may treat very different volumes of water with varying proportions of water from households, rainfall runoff, and other sources. In order to verify that this objective of uniformity is indeed achieved, we studied further the relationship between the WWI and a so-called reference indicator of the virus circulation derived from the incidence rate, which is considered as having a good comparative ability. If the objective of uniformity is reached, we expect this relationship to be the same whichever plant is considered.

To test the achievement of the uniformity objective, we consider the following 3 nested linear mixed effects models of increasing complexity:

- The first one is the simple linear model (Model 0) which corresponds to the case when the homogeneity objective is fully fulfilled:

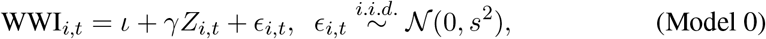

where WWI_*i,t*_ is the WWI value at time t for plant i, *Z*_*i,t*_ is the corresponding reference indicator, *ι* ∈ ℝ, *γ* ∈ ℝ (the intercept and the slope in the linear relation) and *s* ∈ ℝ^+^ (the level of uncertainty of the relation) are parameters to be estimated.
- The second one is a mixed effect model (Model 1) with a random effect on the intercept. It corresponds to the case when the homogeneity target is fulfilled with regard to the multiplicative relation with the reference indicator, but not with regard to the additive relation with the reference indicator:

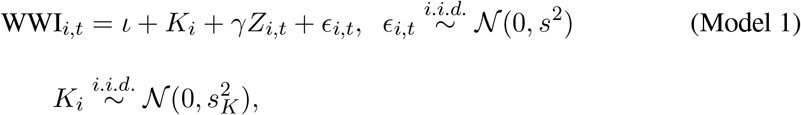

where, in addition to the terms of Model 0, *K*_*i*_ is the intercept random effect for plant *i* and *s*_*K*_ ∈ ℝ^+^ is a parameter to be estimated.
- The third and last one is a mixed effects model with 2 random effects (Model 2). It corresponds to the case when the homogeneity target is not fulfilled with regard to the multiplicative relation nor with regard to the additive relation with the reference indicator:

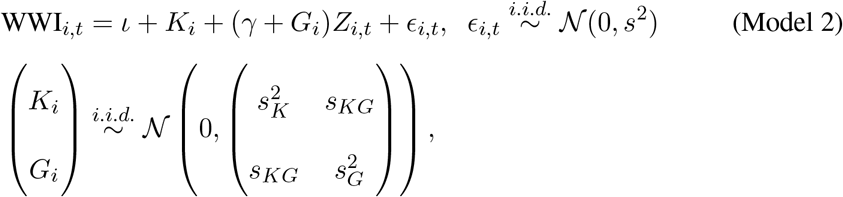

where, in addition to the terms of Model 1, *s*_*KG*_ ∈ ℝ and *s*_*G*_ ∈ ℝ^+^ are parameters to be estimated and *G*_*i*_ is the slope random effect of plant *i*.

In the study that follows, the reference indicator, *Z*, is the logarithm of the incidence rate of the geographic area connected to the treatment plant considered at the same date. In effect, this indicator is considered as a good indicator by the sanitary authorities. The logarithmic transformation makes it possible to find a linear growth like the one obtained for the WWI and thus a comparable curve shape. This reference indicator can be assumed to be universal when it is not affected by public health policies or population movements, for example. We thus restrict the study to the so-called second wave of the epidemic in France excluding main holiday periods, from September the 1^*st*^, 2020 to December the 15^*th*^, 2020.

We estimated a time lag between the two indicators the same way we did in section 3.4, and temporally realigned them accordingly. The focus is on all WWTPs which were analyzed at that time and for which the incidence rate is available for the related municipalities, even though the surveyed populations are not always exactly the same, but considered close enough. To learn the model parameters, we only use the points for which we have measurements at the WWTPs. This notably permits to measure the gain in comparative ability along the successive stages of the WWI construction.

Figure 14 shows the relation between the WWI and the incidence rate in log scale from the full mixed effects model (Model 2). Among the WTTPs considered for the training of the models, one has a stronger negative impact on the comparative ability of the WWI than the others, *Montpellier-Maera*, with an intercept significantly higher than the ones of the other WWTPs, resulting in a potential positive bias. The difference could partly be explained by the fact that the related laboratory only treats this WWTP and two close cities, which complicates the automatic recalibration of this laboratory with regard to the other laboratories as it cannot cover a wide range of the French territory.

**Figure 13:**
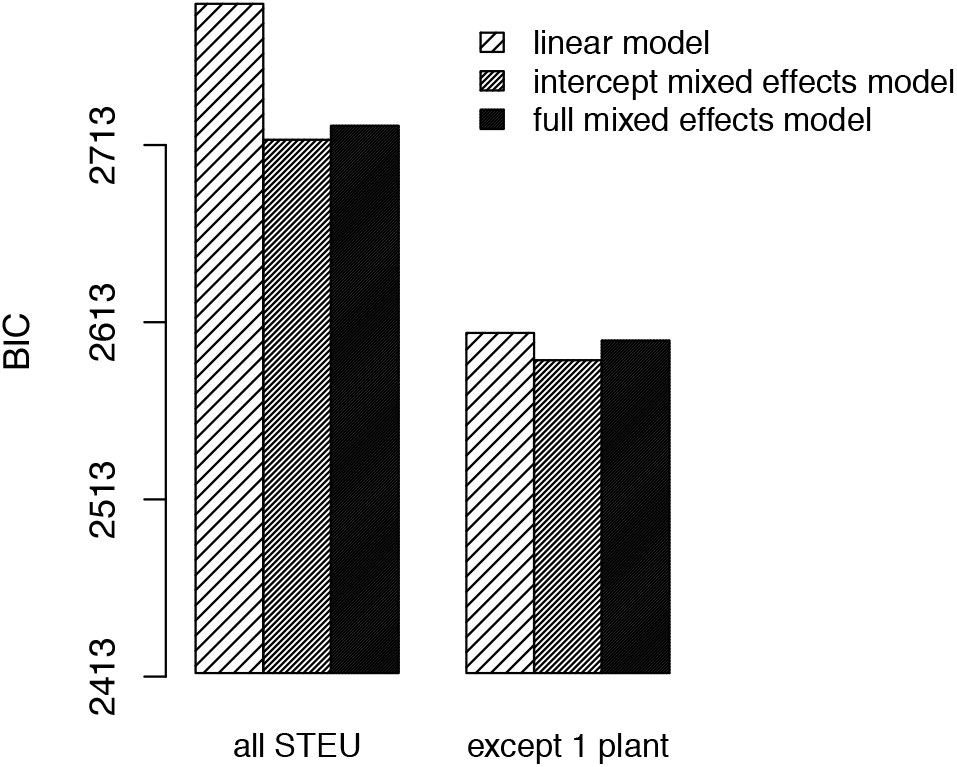
Comparison of Model 2 (full mixed effects model), Model 1 (intercept-only mixed effects model) and Model 0 (simple linear model) according to the Bayesian Information Criterion (BIC) before and after excluding one deviating WWTP (*Montpellier-Maera*). The lower the BIC is, the better the corresponding model is. Model 1 is thus selected while Model 2 is excluded.

**Figure 14:**
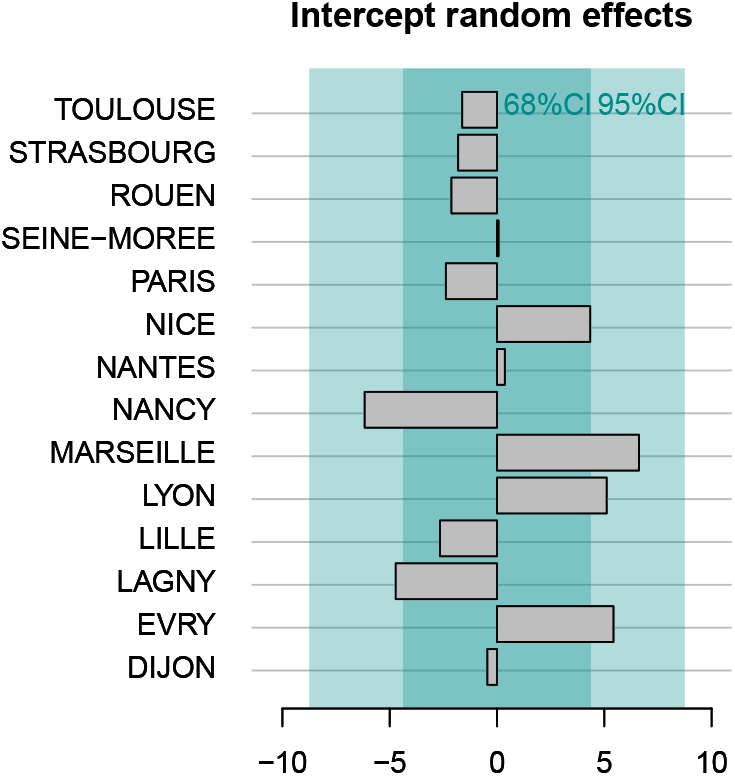
Intercept random effects for Model 1 during the second wave of the epidemic for 14 WWTPs. A positive (resp. negative) intercept effect means the WWI should be lowered (resp. increased) in order to reflect the epidemic state in the same way that the incidence rate does. The deviations at most shortly exceed 5 units of the WWI: for *Nancy, Lagny-sur-Marne* (negative intercept effect), *Marseille, Lyon*, and *Evry* (positive intercept effects) which is acceptable, the WWI typically ranging from -50 to 150. The WWTP of *Paris Seine-Amont* was used for the comparison with the *Grand Paris* incidence rate.

The results of models comparisons according to the BIC^9^ criterion are shown Figure 13. The lower the BIC, the better the performance of the evaluated model. The universal nature of the WWI is validated for the multiplier coefficient (higher performance of Model 1 compared to Model 2). If, in addition, the *Montpellier-Maera* sewage plant is excluded, comparative ability is greatly improved (performance of the mixed-effects models and of the simple linear model are closer), although the difference in performance remains significant and in favor of the intercept mixed-effect model (Model 1).

The (intercept) random effects learned with the selected model (Model 1) after removing the *Montpellier-Maera* WWTP are shown Figure 14. They correspond to the deviation of the WWI of the considered WWTPs from the standard relation between the WWIs and the city incidence rates. A positive (resp. negative) intercept random effect means the WWI should be lowered (resp. increased) in order to reflect the epidemic state in the same way that the incidence rate does. The deviations at most shortly exceed 5 units of the WWI: for *Nancy, Lagny-sur-Marne* (negative intercept effects), *Marseille, Lyon* and *Evry* (positive intercept effects) which is acceptable, the WWI typically ranging from -50 to 150.

Likelihood ratio tests between the nested models show that the comparative ability is improved by each stage of the WWI construction. Indeed, the p-values for the comparison of the mixed effects model on the intercept (Model 1) with the simple linear model (Model 0) (after exclusion of the *Montpellier-Maera* WWTP) strongly increases as we move from the raw data (measurements performed at the WWTP, p-value of 5.10^−34^) to the data accounting for the inlet volumes and de-noised by the previously described smoother (p-value of 9.10^−12^) and to the WWI (p-value of 4.10^−6^).

## 4 Discussion

We have proposed an innovative approach to solve some inherent shortcomings of SARS-CoV-2 analysis in WWTP as a tool to evaluate COVID-19 epidemic. The present algorithm was used in the context of Obepine, a French national surveillance network that is monitoring virus load in 168 WWTPs as of 26th August, 2020. The relevance of WBE^10^ as a decision support tool [29, 30] at the highest political level has been concretely demonstrated in this project. This algorithm allows reducing the measurement noise and taking into account the deviations of quantification between different laboratories. It also makes possible to consider the variations of flow at the inlet of the WWTP, among which the effects of dilutions due to rainfalls, regardless of the size of the WWTP. The signal resulting from this modeling is strongly correlated to the incidence signal in exponential regime, which is consistent with the results of [1, 9, 12, 25, 26, 27, 28]. Outside this regime, the correlation may be weaker, probably because the signal captured by the wastewater analyses is not limited to the detection of virus carriers by massive testing campaigns. Indeed, individual testing is most often restricted to symptomatic and contact cases and may not be representative of virus prevalence in people with no or mild symptoms, notably young people, as previously pointed out [15]. It has indeed been reported that asymptomatic patients may test positive for RT-qPCR in stools [16, 17, 18], thus likely to be detected through wastewater analysis. Moreover, some virus carriers tested negative for RT-qPCR in nasopharyngeal or oropharyngeal swabs, meaning that they would not have been included in the calculation of incidence cases, had they been tested through contact tracing [16, 17].

Based on the data at our disposal, three days sampling seems to be the optimal cost-performance tradeoff to achieve the same kind of results than with an each day sampling process. Although results seem already satisfying for twice a week sampling considering the same criterion and agrees with the conclusions of [21], one could argue that you could still get quite “unlucky” with some two days combinations, whereas this kind of situation would not occur with the three days combinations we studied. Thus, if the budget is not compatible with three days sampling, option 2’, corresponding to biweekly sampling with at least two days without sampling between each sample, might be the best compromise (see Figure 10). It is still important to underline that, even if we were to sample 1000 WWTPs every day of the week, it would only represent 7000 RT-qPCR analyses a week, and give a faithful representation of the epidemic. On the other hand, there were, on average, more than 300 000 tests a week carried out in the single *Île-de-France* region from 13th May, 2020 to 11th June, 2021, according to *Santé Publique France* figures.

Qualitative wise, twice-weekly sampling is still satisfactory, but may lead to the failure to detect some events and affect short-term trends compared to a full week sampling, which is expected as downgrading the sampling frequency reduces the information collected. A bias remains in this subsampling study as sampling was not always done every day of the week at the *Reims* WWTP before November. However, the level of virus circulation did not vary enough between November 2020 and May 2021 to consider a study starting only from November. In particular, this would not have allowed us to account for the fact of detecting none, one or more singular points when the virus becomes quantifiable at a time when the level is generally below the quantification threshold of the analyses (during summer 2020 in the present study). Moreover, we could not try and replicate this subsampling experiment on another WWTP. The same study needs to be replicated on several WWTPs in order to generalize those results with certainty.

The results of lag estimation between the wastewater signal and the incidence rate are in the order of magnitude of a couple days during the exponential phase. Some plants show quite important lags compared to the others, for example *Nancy* WWTP where the WWI lags by 5 days on average and where the intra-experimental variance is more pronounced than in the other plants, or *Paris Seine-Morée* where the WWI is 6 days ahead of the incidence rate signal. Several hypotheses seem plausible to explain these shifts. First, biweekly sampling, although sufficient to capture the dynamics of the epidemic, may induce an additional uncertainty of a few days on the actual peak of excretion in wastewater. Furthermore, the signal captured in wastewaters extends beyond simple reported positive cases. The propensity of populations to test themselves sometimes differs between agglomerations. For two metropolitan areas of similar size, such as *Nancy* and *Mulhouse*, the average rate of testing during the third wave was more than 1.5 times higher in *Nancy*. In municipalities where people test particularly little or more than the average, the indicator is therefore more likely to be ahead or to lag behind the incidence by a few days.

Finally, the good transposition capacity of the WWI from one WWTP to another, relative to what can be observed on the incidence rate signal, is to be considered. Even though it can still be worked upon, our study shows a significant improvement to this property thanks to our smoothing and normalization techniques. It should be noted that the more pronounced deviations in certain plants can have several interpretations, as can the difference between the different lags observed. For example, the incidence rate is only available for the whole of the Aix-Marseille agglomeration, which covers a much larger population than the only plant we monitor in the network in Marseille. The same applies to regional indicators, where the difference in correlation between the two regions could be explained by the deviation in surveyed populations. 28 WWTPs, with a nominal waterflow accounting for around 58% of the regional population, were followed in the *Grand-Est* region while 7 were studied in the *Île-de-France* region (accounting for around 33% of the regional population), leading to a less accurate mesh.

Despite satisfying results, there is still room for improvement. About the inter-laboratory variability assessment, nothing would quite match the possibility to assess the different laboratories on large scale ILA with samples covering a wide range of values in log-scale. Yet, in view of the urgency of the epidemic situation in France from January 2021 and the need to quickly obtain models to help decision-making at the highest political level, the project moved into an action research phase. Monitored sewage plants and analysis laboratories doubled in no less than two months, with analysis reports having to be processed at least once a week. As such ILA results were not available at that time, with some laboratories having no prior history between June 2020 and January 2021, the proposed modelisation was considered as our best option. It shows a great improvement in reducing inter-laboratories variability as shown in Figure 4. Yet, this normalization is not as effective as scaling from ILA results, notably because it is asymmetrical. The problem is that it was not possible to set *C*_*m*_ as a minimum concentration value specific to each laboratory as the true minimum values are censored by quantification and detection thresholds specific to each laboratory. Moreover, *C*_*m*_ was originally designed to be the specific quantification threshold of each laboratory, so that the 0 level would correspond to this quantification threshold for each WWTP. However, one of the laboratory joining late still has a quantification threshold of 40 times the 1000 GU/L limit we are using for *C*_*m*_ as of 19th June, 2021. Using a specifc *C*_*m*_ in the normalization step of the WWI would then have had greatly underestimated the epidemic situation for his related WWTPs. Finally, SARS-CoV-2 circulation level was high in France when we were asked to start communicating our results, hence why we chose a normalization technique that would be more accurate for higher values, yet could still be improved for lower ones.

About the regional indicator, we chose not to use a simple average of the WWI to account for cases where very small WWTP would then have a disproportionate weight in the regional signal. The downside of it is that it accounts less for geographical diversity. For example, if two WWTPs are monitored in a region, with one in the north being really large and one in the south being quite small, the regional WWI will mostly reflect the northern status. An alternative to cope with this problem without extra cost would have been to cluster the clinical signals at city level and associate them with the WWI signals they had a strong correlation with in the same region. Then, the weighted average could have been computed not only with the populations connected to each plant, but with the sum of the populations of the cities which clinical signals had a strong correlation with an WWI. Unfortunately, clinical signals not being openly available at a local level, such a modelisation was not deemed possible.

## 5 Conclusion

The underlying signal in wastewater measurements of SARS-CoV-2 faithfully reflects the dynamics of the epidemic and has the advantage of being unbiased by test availability, willingness of populations to be tested, and population movements. In certain periods, the WWI is also more faithful to the true epidemic situation than the incidence rate, which is obtained as a rolling week average and is therefore very sensitive to holidays (uncharacteristic collapse of the epidemic situation at the peak of the third wave of the pandemic on the incidence rate signal). Moreover, the measurement of this epidemic signal in wastewater proves to be much less costly than massive individual testing. Indeed, it allows obtaining a signal strongly correlated to the more usual epidemic indicators by requiring a single analysis to reflect the average epidemic situation of thousands of people. Finally, this indicator provides an unbiased survey of the infected population, as it also accounts for the contribution of asymptomatic infected persons, which is only partially reflected in the positive test reports, and of unreported infection cases to be recovered. The signal that emerges from these analyses is strongly correlated with the incidence rate and we consider it to be a credible alternative to the latter as its relevance could decline in a few months with the advance of the vaccination campaign and therefore a likely reduction in the quantity of tests carried out to monitor the epidemic.

## Data Availability

The local and regional WWI data are freely available for all WWTPs treated by the Obepine network on the data.gouv.fr public platform. Incidence rate data are partially available in open access for 22 EPCIs and can be found on the same platform.

## Acknowledgements and funding

This work was carried out within the Obepine project, funded by the Ministère de l’Enseignement Supérieur, de la Recherche et de l’Innovation. Financial support was also obtained from CNRS and Sorbonne Université.

We would like to thank *Santé Publique France* for the transmission of specific epidemic data on the watersheds of three wastewater plants used for this study. We would also like to thank Jean-Luc Almayrac from SIAAP for helpful discussions. We would also like to thank Nicolas Benoît from the SACADO unit as well as Sébastien Le Gall and Raphaël Apard for the design, development and maintenance of the data collection and storage platform. We would like to thank the IFREMER for the organization and management of the various ILA. Finally, we would also like to thank the communities of municipalities that agreed on joining the network, the operators that greatly made easier the sampling campaign and all the laboratories that took part in the network.

## Footnotes

1 Wastewater treatment plants

2 Wastewater indicator

3 *Etablissement Public de Coopération Intercommunale* ***(EPCI)***, a French administrative structure that brings together several municipalities in order to exercise some of their common duties.

4 Chemical Oxygen Demand

5 Experimental Data Quality and Precision Indicator

6 *Ministère de la Transition écologique et solidaire*

7 In practice, *ℓ* can vary from one day to another, for instance if one works on quantities that correspond to the multiplication of concentrations (with a detection limit) by a fluctuating volume. This can be taken into account within our method with no additional cost.

8 Inter-laboratory assays

9 Bayesian Information Criterion

10 Wastewater-based epidemiology

